# SPLIT: Safety Prioritization for Long COVID Drug Repurposing via a Causal Integrated Targeting Framework

**DOI:** 10.64898/2026.04.12.26350701

**Authors:** Sindy Pinero, Xiaomei Li, Lin Liu, Jiuyong Li, Sang Hong Lee, Thuc Duy Le

## Abstract

Long COVID affects millions of people worldwide, yet no disease-modifying treatment has been approved, and existing interventions have shown only modest and inconsistent benefits. A key reason for this limited progress is that current computational drug repurposing pipelines do not match well with the clinical reality of Long COVID. These patients often have persistent, multi-systemic symptoms and may already be taking multiple medications, making treatment safety a primary concern. However, most repurposing workflows still treat safety as a downstream filter and rely on disease-associated targets rather than causal drivers. They also assume that the findings of one analysis would generalize across the diverse presentations of Long COVID. We introduce SPLIT, a safety-first repurposing framework that addresses these limitations. SPLIT prioritizes safety at the start of the candidate evaluation, integrates complementary causal inference strategies to identify likely driver genes, and uses a counterfactual substitution design to compare drugs within specific cohort contexts. When applied to cognitive and respiratory Long COVID cohorts, SPLIT revealed three main findings. First, drugs with similar predicted efficacy could have very different predicted safety profiles. Second, the drugs flagged as unfavorable were often different between the two cohorts, showing that drug prioritization is phenotype-specific. Third, SPLIT flagged 18 drugs currently under active investigation in Long COVID trials as having unfavorable predicted profiles. SPLIT provides a practical framework to identify safer, more context-appropriate candidates earlier in the process, supporting more targeted and better-tolerated treatment strategies for Long COVID.

## INTRODUCTION

Long COVID has become a significant chronic health problem following the COVID-19 pandemic, affecting millions of people worldwide^1–4^. Despite intense clinical and computational efforts, no disease-modifying treatment has been approved^5^, and the available interventions have shown only modest and inconsistent benefits^6–10^. This limited progress suggests that current drug repurposing strategies may not be well aligned with the clinical reality of Long COVID.

A major challenge is that Long COVID is a chronic, multisystemic condition. Patients may experience persistent symptoms in the neurological, respiratory, cardiovascular, and metabolic domains, often while using multiple medications^1,4,11,12^. In this setting, the safety and tolerability of treatment are especially important. A drug that adds substantial adverse effects is unlikely to be acceptable for long-term treatment, even if it appears to be mechanistically promising. However, most computational repurposing pipelines still prioritize efficacy-related or mechanistic signals first and consider safety only after candidate drugs have been selected^13–16^. For Long COVID, this ordering is not ideal.

A second limitation is the way candidate targets are often chosen. Many repurposing methods rely on genes or proteins that are associated with disease through changes in expression, network proximity, or other correlation-based signals^17–19^. These approaches can identify biologically relevant markers, but do not distinguish between genes that simply reflect disease processes and those that drive them. As a result, candidate drugs may be directed towards downstream consequences rather than to meaningful intervention points^17–19^. For a complex condition such as Long COVID, this can weaken the translational value of computational predictions.

A third limitation is the assumption that one repurposing result will generalize across all Long COVID patients. Long COVID is highly heterogeneous, with different clinical presentations and likely different underlying mechanisms^1,3^. Patients with primarily cognitive symptoms may differ substantially from those with primarily respiratory symptoms in baseline biology, comorbidities, and treatment risk-benefit profiles. A drug that appears suitable in one context may be less suitable in another. This means that phenotype-specific evaluation is likely to be important for meaningful candidate prioritization.

To address these limitations, we developed **SPLIT** (**S**afety **P**rioritization for **L**ong COVID Drug Repurposing via a Causal **I**ntegrated **T**argeting Framework). SPLIT is a safety-first computational framework that prioritizes safety at the beginning of candidate evaluation, identifies putative driver genes using three complementary causal inference strategies: Transcriptome-Wide Mendelian Randomization (TWMR)^20^, Control Theory (CT)^21^, and Differential Causal Effects (DCE)^22^, and compares candidate drugs within fixed cohort contexts using a counterfactual substitution design. Rather than relying solely on disease associations, SPLIT aims to repurpose targets with stronger causal relevance and to evaluate drugs that better reflect the heterogeneity of Long COVID.

We applied SPLIT to cognitive and respiratory Long COVID cohorts and found three main results. First, drugs with similar predicted efficacy can differ markedly in predicted safety, suggesting that efficacy alone is not sufficient to distinguish between candidates. Second, drugs flagged as unfavorable often differed between cohorts, indicating that drug prioritization is strongly phenotype-specific. Third, SPLIT identified 18 drugs currently under active investigation in Long COVID trials as having unfavorable predicted profiles. By identifying safer candidates, more context-appropriate, and earlier, SPLIT provides a practical framework to support more targeted and better-tolerated treatment strategies for Long COVID.

## RESULTS

### Safety Prioritization Framework for Long COVID Drug Repurposing

We developed SPLIT, a computational framework that combines multi-omics data, causal gene inference, and clinical knowledge graph modeling to rank Long COVID drug candidates by safety first, before any efficacy considerations are applied.

The framework runs in five stages (Figure 1):

1. **Long COVID (LC) Causal Genes (Panel A):** We identified the likely Long COVID driver genes using three complementary causal inference strategies: Mendelian Randomization (MR), Control Theory (CT) and Differential Causal Effects (DCE) by integrating expression Quantitative Trait Loci (eQTL), Genome-Wide Association Study (GWAS), Ribonucleic Acid sequencing (RNA-seq) gene expression, Protein–Protein Interaction (PPI) networks, and Kyoto Encyclopedia of Genes and Genomes (KEGG) pathway data.
2. **Integrative Method (Panel B):** We combined causal evidence to obtain consensus driver genes for Long COVID, linked them to candidate drugs through drug–gene mapping, and generated drug repurposing predictions using PlaNet^23^.
3. **Validation and Scoring (Panel C):** We validated candidate drugs in independent Long COVID cohorts, used PlaNet^23^ to predict safety (S), adverse event burden (AE) and comparative efficacy (E) for each candidate, and developed a composite deprioritisation ranking score that integrates S, AE and E to identify drug candidates with high-risk for deprioritization.
4. **Downstream Analysis (Panel D):** We predicted S/AE/E probabilities across different Long COVID cohorts and enriched the top deprioritized candidate drugs with wet-lab and pharmacological evidence from public databases.
5. **Clinical Outputs (Panel E):** We translated the results into practical outputs, producing lists of deprioritized candidates that should be avoided for clinical translation.

**Figure 1:**
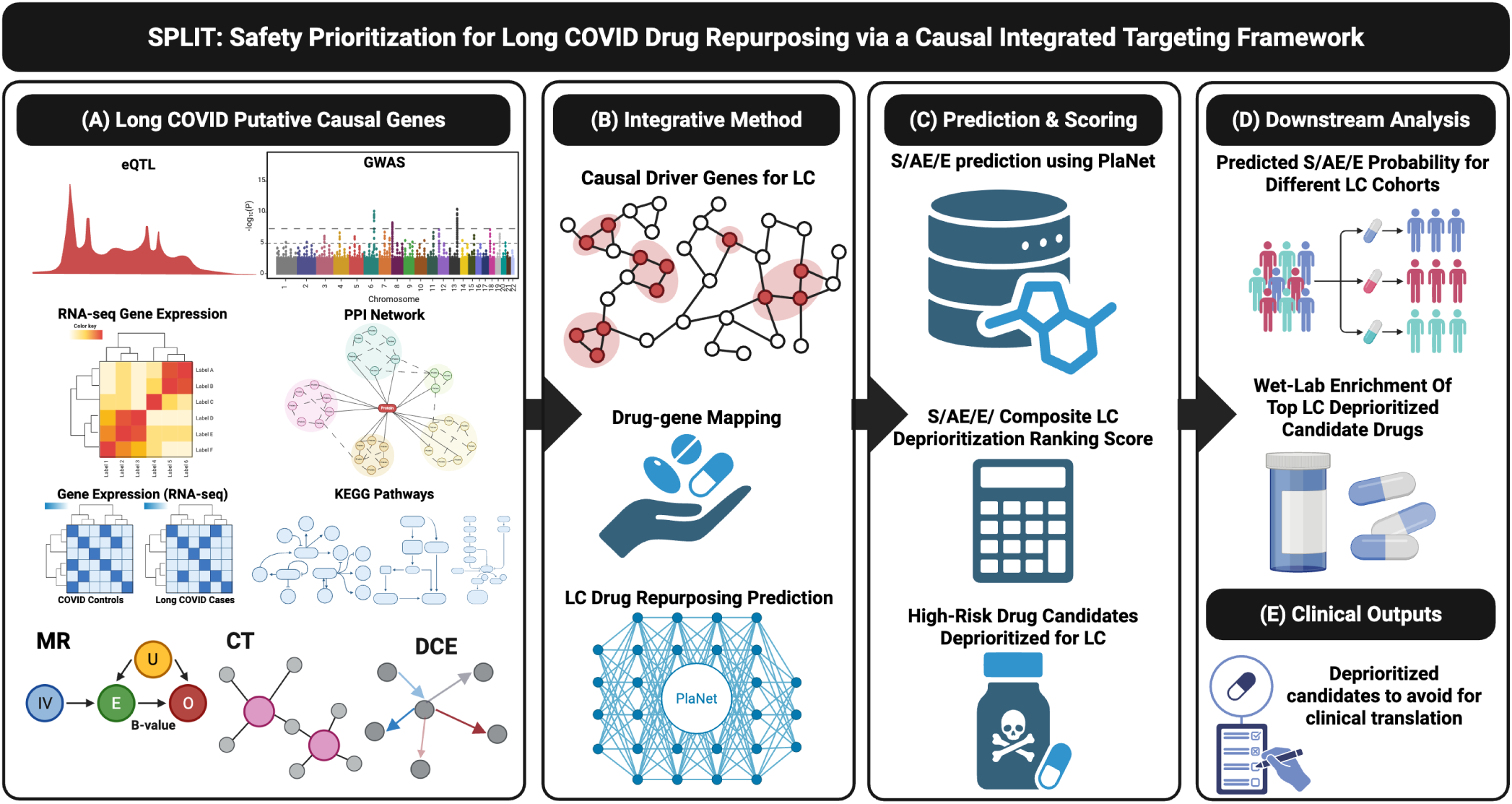
Safety Prioritization Framework for Long COVID Drug Repurposing (SPLIT). (A) Multi-omics input data including eQTL, GWAS, RNA-seq, and PPI networks. (B) Three complementary causal inference approaches, TWMR, CT, and DCE, are integrated to produce a unified Long COVID driver gene set. (C) Driver genes are mapped to drugs, and S, AE, and E are predicted for each candidate within a fixed cohort context via counterfactual substitution. (D) Composite scoring, deprioritization ranking, and pharmacological enrichment. (E) Clinical outputs: avoid lists and shortlists for trial design. Created in BioRender. Pinero, S. (2025) License. **Abbreviations:** eQTL, expression Quantitative Trait Locus; GWAS, Genome-Wide Association Studies; RNA-seq, RNA sequencing; PPI, protein-protein interaction; TWMR, Transcriptome-Wide Mendelian Randomization; CT, Control Theory; DCE, Differential Causal Effects; S, predicted safety score; AE, adverse event burden; E, comparative efficacy.

The input data include Genome-Wide Association Studies (GWAS), quantitative trait locus cis-expression data (cis-eQTL), RNA sequencing (RNA-seq) from Long COVID cases and controls, and a protein-protein interaction (PPI) network (Figure 1A). These data feed into three causal inference methods, each designed to capture a different aspect of driver biology: Transcriptome Wide Mendelian Randomization (TWMR), which focuses on genes whose genetically predicted expression causally contributes to Long COVID risk; Control Theory (CT), which identifies regulatory nodes in the PPI network that are essential for network function; and Differential Causal Effects (DCE), which measures how pathway activity changes specifically in Long COVID patients compared to healthy controls (Figure 1B).

When we applied SPLIT to the harmonized Long COVID multi-omics data, the three causal inference methods identified a total of 1,725 likely driver genes (Figure 3A; Table 1). CT contributed the largest set (1,641 genes), which reflects the scale of the PPI network, while TWMR (50 genes) and DCE (45 genes) provided more targeted, complementary evidence. Eleven genes were supported by two or more methods, making them the causal drivers with the highest confidence (Table 1). These include immune regulators (*STAT3*, *HIF1A*), cellular stress response genes (*TP53*, *MYC*), signaling components (*POMC*, *LEF1*), and the DNA repair factor *ERCC3*. The complete gene list with annotations is provided in ST S1.

**Table 1:** Long COVID causal driver genes supported by two or more causal inference methods. CT, Control Theory; MR, Mendelian Randomization; DCE, Differential Causal Effects. Complete annotations in ST S1.

| Gene | Supporting methods | Functional category |
| --- | --- | --- |
| *ERCC3* | CT; MR | DNA repair |
| *HIF1A* | CT; DCE | Hypoxia / immune regulation |
| *IMMT* | CT; MR | Mitochondrial structure |
| *LEF1* | CT; DCE | Wnt signaling |
| *MED19* | CT; MR | Transcription (Mediator complex) |
| *MYC* | CT; DCE | Cellular stress / proliferation |
| *NDUFA6* | CT; MR | Mitochondrial complex I |
| *PNPO* | DCE; MR | Vitamin B<sub>6</sub> metabolism |
| *POMC* | CT; DCE | Neuroendocrine signaling |
| *STAT3* | CT; DCE | Immune regulation / JAK-STAT |
| *TP53* | CT; DCE | Cell cycle / DNA damage |

When we mapped these 1,725 genes to five curated drug-target databases (DrugBank, ChEMBL, TTD, DrugCentral, GtoPdb), we found 19,172 unique drugs with at least one causal gene target. To ensure that each candidate had a meaningful mechanistic coverage, we restricted the screening panel to drugs that target at least 11 causal genes. This threshold was supported by four lines of evidence, including hypergeometric enrichment analysis at which 100% of retained drugs show a statistically non-random overlap with the causal gene network (FDR *<* 0.05), distributional analysis, safety profile comparisons across thresholds, and per-method panel contribution tests; the full derivation and sensitivity analysis are provided in ST S7. This produced 1,639 candidates per cohort (median: 35 targets per drug; range: 11–210). The drugs with the broadest coverage of causal genes were predominantly oncology kinase inhibitors, such as imatinib (210 targets) and sirolimus (206 targets). Complete drug-gene mapping and drug target annotations are provided in ST S2.

### Drugs to Avoid for Long COVID Patients

We applied SPLIT to two Long COVID clinical trial cohorts representing different patient populations: a cognitive cohort (N = 72) and a respiratory cohort (N = 32) (Table 5 in the Methods Section). For each cohort, we predicted S, E, and AE for all 1,639 candidate drugs and ranked them using a composite score that combines all three dimensions (ST S21). At the default threshold (worst 10%), 163 drugs were flagged for deprioritization in each cohort.

The top 10 deprioritized drugs from each cohort showed fundamentally different failure patterns (Table 2). In the cognitive cohort, deprioritized drugs had consistently low predicted safety (S ranging from 0.123 to 0.404), while predicted efficacy clustered close to placebo (≈0.52). Drug classes included JAK inhibitors (upadacitinib), antimicrobials (secnidazole, pivampicillin, fidaxomicin, ethionamide), corticosteroids (fluticasone), an SGLT2 inhibitor (empagliflozin), and a phenothiazine (thiethylperazine). In the respiratory cohort, the avoid list was dominated by cancer drugs (pazopanib, azacitidine, carboplatin, paclitaxel, decitabine, temozolomide, and temsirolimus), all with E values below 0.05, indicating that the placebo was predicted to be superior. This reflects the mechanistic mismatch between cytotoxic agents and respiratory recovery after viral infection. Selexipag, despite its FDA-approved indication for pulmonary arterial hypertension, was also deprioritized due to a high AE burden (7.9) and moderate predicted efficacy (0.112), consistent with the tolerability challenges documented in the GRIPHON trial^24^.

**Table 2:** Top 10 deprioritized drugs from the primary composite avoid list for both cohorts (q10). The cognitive cohort is driven by low S (84.7% of q10 drugs have S *<* 0.3); the respiratory cohort by low E (100% of q10 drugs have E *<* 0.5) and high AE burden (98.2% have AE *>* 2). Full lists in ST S9 and ST S17.

| NCT04809974 (Cognitive) |  |  |  |  |  | NCT04880161 (Respiratory) |  |  |  |  |  |
| --- | --- | --- | --- | --- | --- | --- | --- | --- | --- | --- | --- |
| R | Drug | C | S | E | AE | R | Drug | C | S | E | AE |
| 1 | Upadacitinib | 0.159 | 0.241 | 0.515 | 2.3 | 1 | Pazopanib | 0.147 | 0.380 | 0.024 | 7.4 |
| 2 | Secnidazole | 0.164 | 0.124 | 0.516 | 1.1 | 2 | Azacitidine | 0.156 | 0.370 | 0.047 | 6.8 |
| 3 | Fluticasone P. | 0.185 | 0.127 | 0.516 | 0.8 | 3 | Carboplatin | 0.165 | 0.371 | 0.042 | 4.7 |
| 4 | Pivampicillin | 0.188 | 0.320 | 0.514 | 2.2 | 4 | Paclitaxel | 0.168 | 0.389 | 0.019 | 4.4 |
| 5 | Empagliflozin | 0.191 | 0.183 | 0.517 | 1.7 | 5 | Decitabine | 0.169 | 0.387 | 0.045 | 6.6 |
| 6 | Fidaxomicin | 0.191 | 0.200 | 0.517 | 2.0 | 6 | Selexipag | 0.172 | 0.342 | 0.112 | 7.9 |
| 7 | Ethionamide | 0.201 | 0.196 | 0.517 | 1.9 | 7 | Temozolomide | 0.175 | 0.401 | 0.014 | 4.3 |
| 8 | Bithionol | 0.204 | 0.299 | 0.515 | 2.6 | 8 | Donepezil | 0.178 | 0.279 | 0.105 | 2.3 |
| 9 | Thiethylperazine | 0.205 | 0.219 | 0.517 | 2.0 | 9 | Temsirolimus | 0.178 | 0.425 | 0.024 | 8.7 |
| 10 | Tavaborole | 0.210 | 0.168 | 0.519 | 2.3 | 10 | Permethrin | 0.179 | 0.298 | 0.076 | 2.2 |

Abbreviations: R, rank; C, composite score; S, safety; E, efficacy; AE, AE burden; P., propionate.

In addition, SPLIT identified 18 drugs currently under investigation in Long COVID clinical trials as candidates for deprioritization (Table 3; Figure 2). These include metformin, tested in the COVID-OUT phase 3 prevention trial^25^ and nirmatrelvir/ritonavir, tested in the STOP-PASC treatment trial^26^. The JAK inhibitors baricitinib and upadacitinib were deprioritized due to immuno-suppressive risk and FDA black box warnings, but both advanced to phase 3 in the Long COVID trials^27,28^: baricitinib was marked in both cohorts (cognitive S=0.275; respiratory E=0.058) but continues in the REVERSE-LC trial. Symptomatic management agents also appear on the avoid lists, including ivabradine for POTS/dysautonomia^29,30^, modafinil for fatigue on the RECOVER-ENERGIZE platform^31^, and low-dose naltrexone based on observational reports^32,33^. Anticoagulant and antiplatelet combinations (apixaban, clopidogrel, acetylsalicylic acid) reflect the thromboinflammatory hypothesis tested in triple therapy regimens^34^. We emphasize that being on an avoid list does not necessarily rule out clinical investigation: the mechanistic rationale and the specific patient context may justify testing higher-risk agents in selected subgroups, and all predictions require prospective validation. However, these findings highlight candidates that warrant closer safety monitoring and careful reassessment of the benefit-risk balance in ongoing and planned trials.

**Figure 2:**
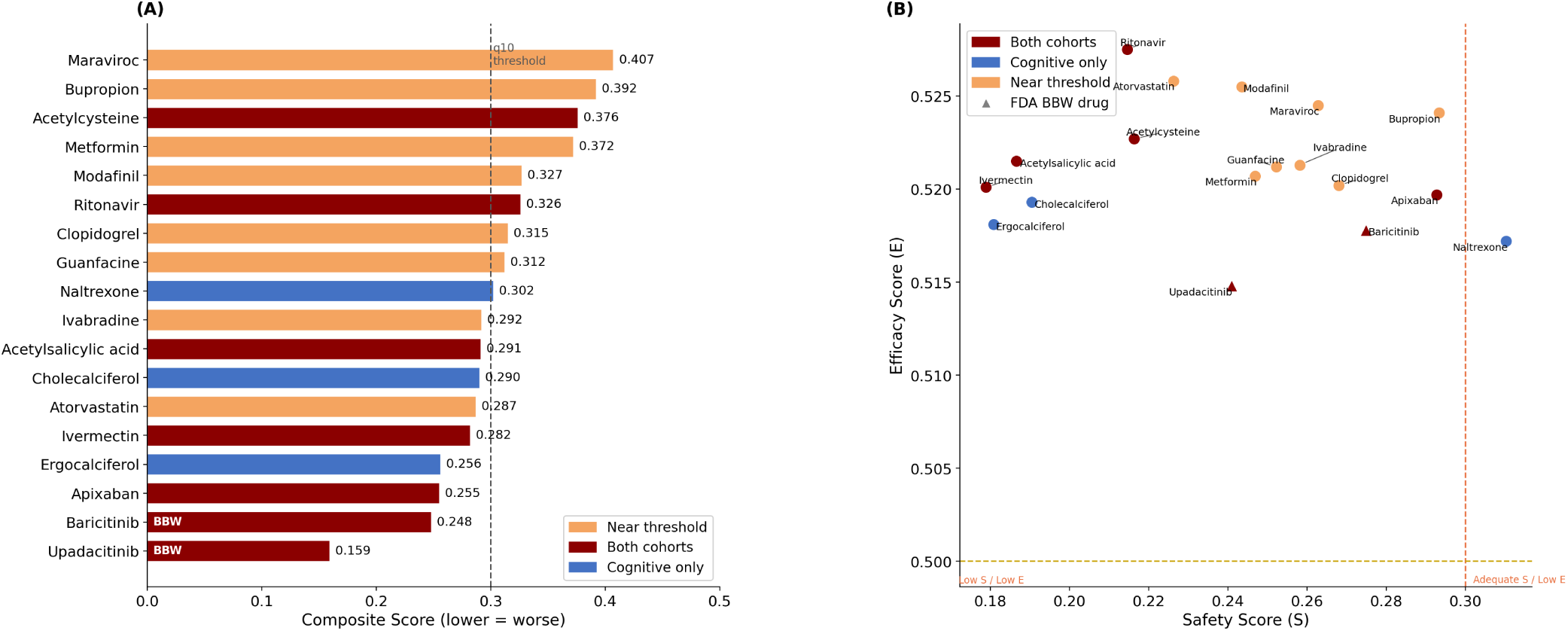
Eighteen active Long COVID trial drugs flagged for deprioritization. (A) Horizontal bars show the worst-case composite score across cohorts (lower = worse). Colors indicate cohort of primary flagging: dark red, both cohorts; blue, cognitive only; red/orange, respiratory only or near threshold. Dashed vertical line: q10 threshold. (B) S versus E scatter in the cognitive cohort for the 18 flagged drugs. Colors indicate which cohort(s) flagged the drug; triangles, FDA BBW drugs. Dashed lines: S=0.3 (vertical), E=0.5 (horizontal). Drugs in the upper-left quadrant (low S, acceptable E) represent safety-driven flags; drugs in the lower half (E *<* 0.5) represent efficacy-driven flags.

**Table 3:** Drugs on the SPLIT avoid lists currently under investigation for Long COVID. Study types: RCT, randomized controlled trial; Reg, registered trial; Clin, clinical report. Avoid reason: primary flag trigger. Literature references in ST S20.

| Drug | Cohort | Study | Mechanism/Target | Avoid reason | Cite |
| --- | --- | --- | --- | --- | --- |
| Acetylcysteine | Both | Clin | Antioxidant; glutathione | Δ worse vs Ampion (q10) | 35 |
| Acetylsalicylic acid | Both | Clin | COX-1 antiplatelet | Δ worse; low S (q10) | 34 |
| Apixaban | Both | Clin | Factor Xa inhibitor | Very low E; high AE (q10) | 34 |
| Atorvastatin | Resp | Reg RCT | Statin; pleiotropic | Low E (q20) | 36 |
| Baricitinib | Both | Reg Ph3 | JAK1/2 inhibitor | High AE; BBW (q10) | 27 |
| Bupropion | Resp | Clin | NDRI antidepressant | Low E (q20) | 37 |
| Cholecalciferol | Both | RCT | Vitamin D | Low S (q10) | 38 |
| Clopidogrel | Resp | Clin | P2Y12 inhibitor | Low E (q20) | 34 |
| Ergocalciferol | Both | RCT | Vitamin D | Low S (q05) | 39 |
| Guanfacine | Resp | Clin | α2A-adrenergic agonist | Low E (q20) | 35 |
| Ivabradine | Resp | Reg RCT | I<sub>f</sub>-channel blocker | Low E (q20) | 29 |
| Ivermectin | Both | Ph3 RCT | Antiparasitic; host-directed | Low S (q10) | 25 |
| Maraviroc | Resp | Reg RCT | CCR5 antagonist | Low E (q20) | 36 |
| Metformin | Resp | Ph3 RCT | Metabolic/AMPK | Low E (q20) | 25 |
| Modafinil | Resp | Reg | Wake-promoting; DAT | Low E (q20) | 31 |
| Naltrexone | Cog | Clin | Opioid receptor antagonist | Δ worse vs Niagen (q10) | 32 |
| Ritonavir | Both | RCT | Protease inhibitor (NMV/r) | Low E (q20) | 26 |
| Upadacitinib | Both | Reg Ph3 | JAK1 inhibitor | Low S; high AE; BBW (q05) | 28 |

**Abbreviations:** Cog, NCT04809974; Resp, NCT04880161; Ph3, Phase 3; BBW, FDA black box warning.

### Drugs with Similar Efficacy Have Different Safety Levels

A key finding from our full drug panel is that similar predicted efficacy does not mean similar safety. Figure 3C illustrates this directly through four counterfactual comparisons: Niagen, Leronlimab, Vortioxetine, and Ritonavir all produce similar E values (0.519–0.527) within the same fixed cohort context, but their predicted S and AE burdens are very different. This shows that efficacy alone is not enough to distinguish between candidates: without an explicit safety evaluation, drugs with very different risk profiles appear interchangeable.

**Figure 3:**
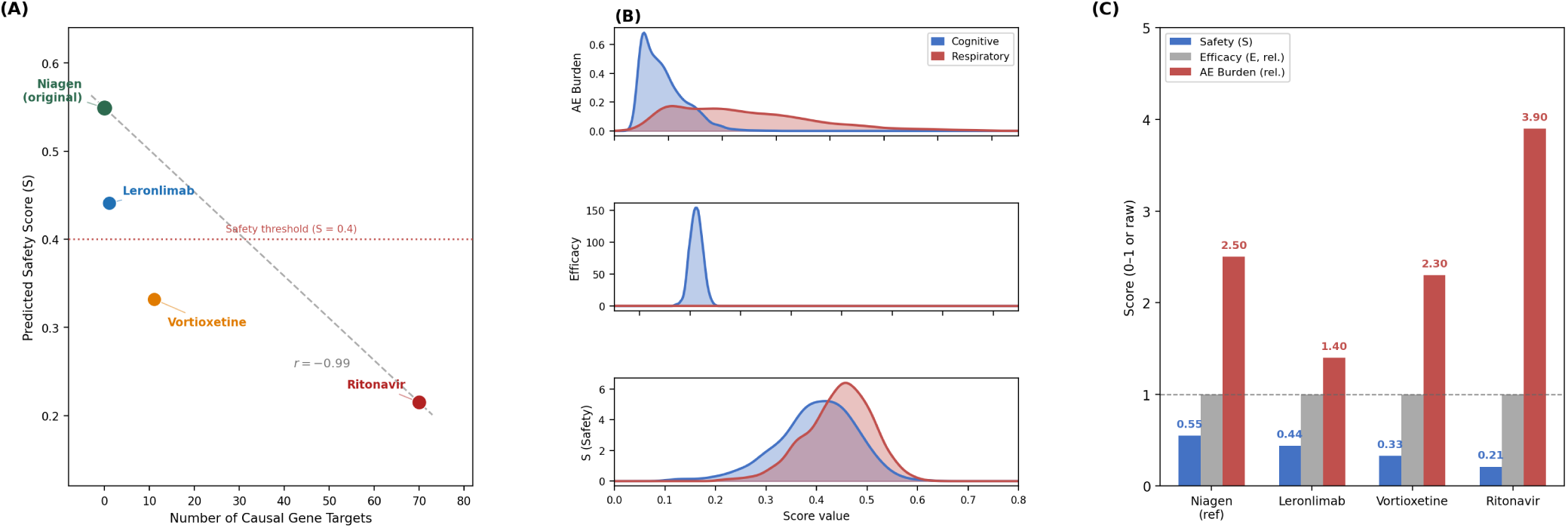
Safety trade-off, score distributions, and counterfactual risk profiles. (A) Predicted S decreases as causal gene coverage increases (*r* = −0.99) across four counterfactual scenarios in the template cohort. Dotted line: S=0.4 threshold. (B) Kernel density distributions of S, E, and AE burden for the full 1,639-drug panel across both cohorts (cognitive and respiratory). E is compressed in the cognitive cohort (range 0.514 to 0.531) but widely spread in the respiratory cohort (0.009 to 0.635). (C) Relative scores for each counterfactual drug: S ranges from 0.21 to 0.55 while E is essentially flat across all four candidates, showing that safety, not efficacy, drives candidate differentiation within a fixed cohort context. **Abbreviations:** S, predicted safety score; E, predicted comparative efficacy score; AE, adverse event burden score.

The underlying reason for this difference is the breadth of drug mechanisms. The relationship between the number of causal gene targets and predicted S was strong and inverse (*r* = −0.99; Figure 3B). Ritonavir, which targets 70 causal genes, had the lowest predicted S (0.215). Vortioxetine (11 targets) showed an intermediate S (0.332), and Leronlimab (1 target) had the highest S among these drugs (0.441). Niagen, a dietary supplement with an established tolerability profile^40^ and no direct causal gene targets, had the highest overall S (0.549). The predicted adverse event profiles matched well-known drug-class signatures: nausea (23.4%) with the serotonergic agent Vortioxetine; diarrhea (19.2%) and skin reactions with the CYP3A4 inhibitor Ritonavir; and hypertensive disease as a recurring background signal across Niagen and Leronlimab, consistent with cardiovascular involvement in Long COVID patients. Together, these results show that drugs acting on fewer, more selective targets tend to have safer predicted profiles, and that safety, rather than efficacy, is the main axis of candidate differentiation within a fixed clinical context.

### Safety Levels of Drugs Are Different Among Long COVID Subtypes

When we compared the avoid lists between cohorts, we found that deprioritization worked through fundamentally different mechanisms in cognitive versus respiratory Long COVID (Table 4; Figure 5). This difference is visible in the score distributions throughout the panel (Figure 4): while the S distributions differ in absolute level between the cohorts but overlap substantially, the E distributions are qualitatively distinct. In the cognitive cohort, E values cluster tightly around 0.52 with very little spread. In the respiratory cohort, E ranges from nearly zero to 0.64. The predicted AE burden is approximately twice that in the respiratory cohort throughout the panel. These distributional differences do not arise from threshold choice; they reflect how differently the two cohort contexts rank the same set of drug candidates.

**Figure 4:**
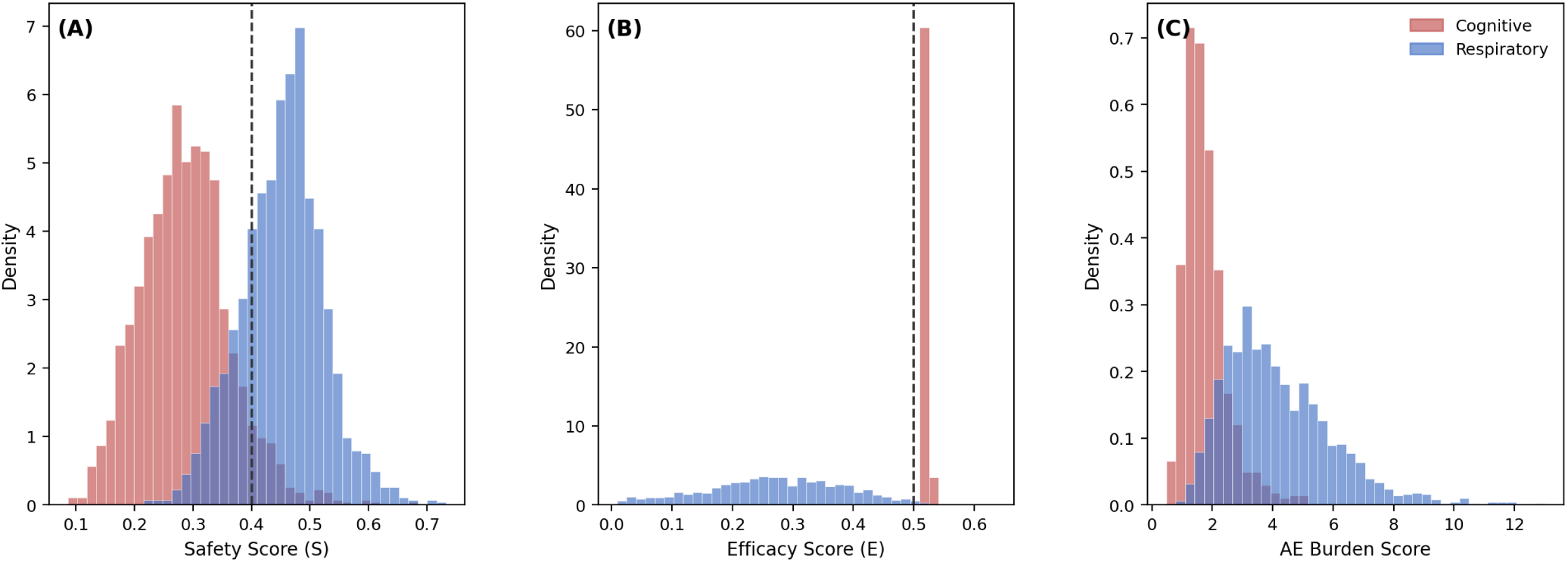
Complete panel score distributions across cohorts. (A) Safety distributions. The cognitive cohort (red) is shifted to lower values (mean 0.286) compared to the respiratory cohort (blue; mean 0.452), with the dashed line at S=0.4 indicating that only 7.0% of cognitive drugs and 78.0% of respiratory drugs exceed the safety threshold. (B) Efficacy distributions. The cognitive cohort is compressed into a narrow spike near 0.52 (range 0.514 to 0.531; spread 0.017), while the respiratory cohort spans the full range from near zero to 0.64 (spread 0.626, 37× larger). This 37-fold difference in E discrimination explains why deprioritization is S-driven in the cognitive cohort and E-driven in the respiratory cohort. (C) AE burden distributions. Both cohorts are right-skewed; the respiratory cohort shows a heavier tail extending to AE *>* 10, reflecting approximately 2× higher burden at the extremes relative to the cognitive cohort. Dashed vertical lines: S=0.4 (left panel) and E=0.5 (center panel). **Abbreviations:** S, predicted safety score; E, predicted comparative efficacy score; AE, adverse event burden score.

**Figure 5:**
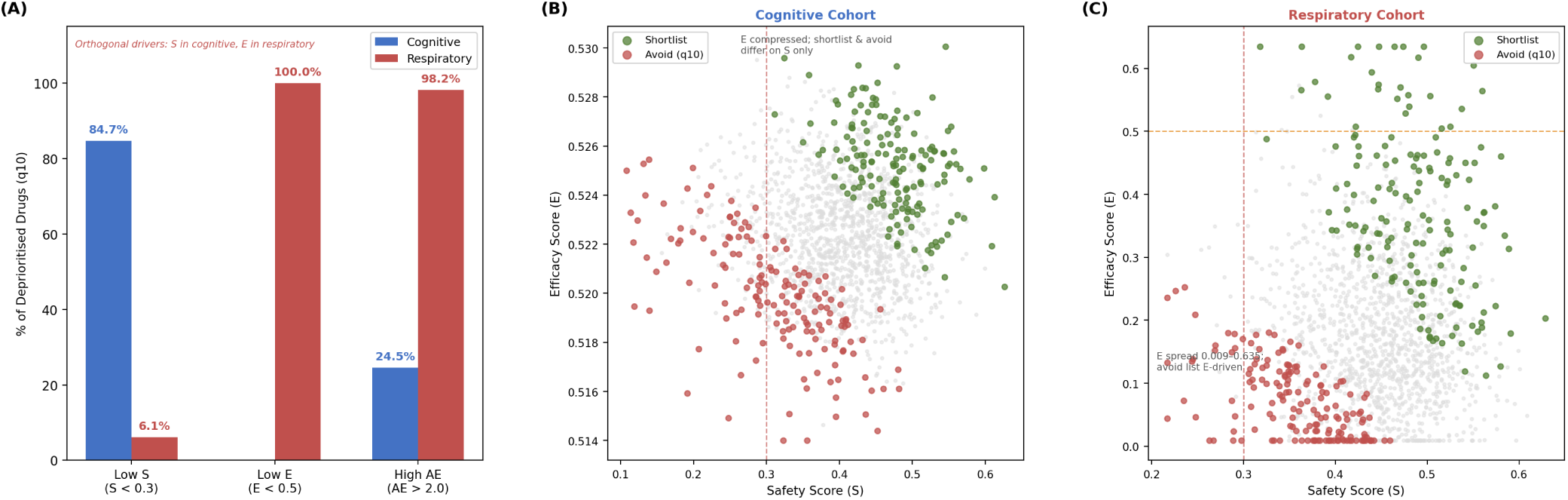
Deprioritization mechanisms differ between cohorts. (A) Proportion of deprioritized drugs (q10) meeting each failure criterion. The cognitive cohort is dominated by low S (84.7%); the respiratory cohort by low E (100%) and high AE burden (98.2%). (B) S versus E scatter for the cognitive cohort. E is compressed near 0.52; shortlisted (dark green) and deprioritized (red) drugs are separated on S only. (C) S versus E scatter for the respiratory cohort. E ranges from 0.009 to 0.635; the deprioritized cluster occupies the low-E region regardless of S. Dashed lines: S=0.3 (vertical), E=0.5 (horizontal). **Abbreviations:** S, predicted safety score; E, predicted comparative efficacy score; AE, adverse event burden score.

**Table 4:** Comparison of deprioritization patterns across cohorts. The two cohorts exhibit distinct dominant drivers: low S in the cognitive cohort versus low E in the respiratory cohort. The 37-fold difference in E spread explains this asymmetry. Only 11% of deprioritized drugs are shared. **Abbreviations:** S, predicted safety score; E, predicted comparative efficacy score; AE, adverse event burden score.

| Metric | Cognitive Cohort | Respiratory Cohort |
| --- | --- | --- |
| **Score discrimination across the full panel** |  |  |
| E spread | 0.017 (minimal) | 0.626 (37× larger) |
| S range | 0.086–0.682 | 0.217–0.733 |
| AE burden range | 0.5–6.1 | 0.9–13.0 ($\approx 2\times$ higher) |
| **Dominant deprioritization driver (q10)** |  |  |
| Dominant signal | Low S ($S < 0.3$) | Low E ($E < 0.5$) |
| % with low S ($< 0.3$) | 84.7% | 6.1% |
| % with low E ($< 0.5$) | 0.0% | 100.0% |
| % with high AE ($> 2$) | 24.5% | 98.2% |
| **Cross-cohort overlap** |  |  |
| Shared deprioritized | 18 drugs (11.0%) |  |
| Unique to each cohort | 145 drugs | 145 drugs |

Of the 163 drugs deprioritized in each cohort, only 18 (11.0%) appeared on both avoid lists. The failure modes were different: in the cognitive cohort, 84.7% of the deprioritized drugs had low S (S *<* 0.3) and none had low E, while in the respiratory cohort, all deprioritized drugs had low E and only 6.1% had low S (Figure 5A). This asymmetry is explained by the 37-fold difference in E discrimination: E ranged from 0.514 to 0.531 in the cognitive panel but from 0.009 to 0.635 in the respiratory panel (Figure 5B and C). When E does not separate candidates, S becomes the main determinant of differentiation. When E varies widely, it drives deprioritization.

The predicted AE burden was approximately 2.4× higher in the respiratory cohort (mean 4.14 vs. 1.76 in the cognitive cohort) and reached 10.4 at the extreme, compared to 3.6 in the cognitive cohort, even for the same drug classes. This suggests that the respiratory Long COVID phenotype amplifies the predicted toxicity, possibly due to baseline airway inflammation and pre-existing lung involvement. Oncology kinase inhibitors accounted for the majority of drugs that failed multiple thresholds simultaneously in both cohorts (≈70% of drugs flagged), but with clearly different profiles: AE values of 9.5–13.0 in the respiratory cohort versus 3.7–5.5 in the cognitive cohort, and E values of 0.03–0.05 versus ≈0.52, respectively.

The 18 drugs deprioritized in both cohorts represent candidates with consistently unfavorable profiles, regardless of the Long COVID phenotype, although they failed through different mechanisms in each cohort. All showed an improvement in S in the respiratory cohort (ΔS=+0.10 to +0.23; mean +0.16) but a reduction in E (ΔE = −0.26 to −0.46; mean −0.37). The fact that they were flagged regardless of which failure mode dominated suggests that their suitability reflects the intrinsic properties of the drugs themselves, rather than an artifact of the cohort context.

Looking through the entire drug panel, we identified 16 candidates who ranked in the top 10% by composite score in both cohorts. These cross-phenotype shortlisted drugs showed consistent improvements in S when moving from the cognitive to respiratory cohort (ΔS=+0.098 to +0.230) while maintaining useful E values (Figure 6A). We also identified 26 drugs with conflicting profiles between cohorts, appearing on the avoid list in one cohort but on the shortlist in the other, which highlights the practical consequences of phenotype-specific evaluation (Figure 6B). Complete data on cross-cohort shifts and conflict drug lists are provided in ST S9 and ST S17.

**Figure 6:**
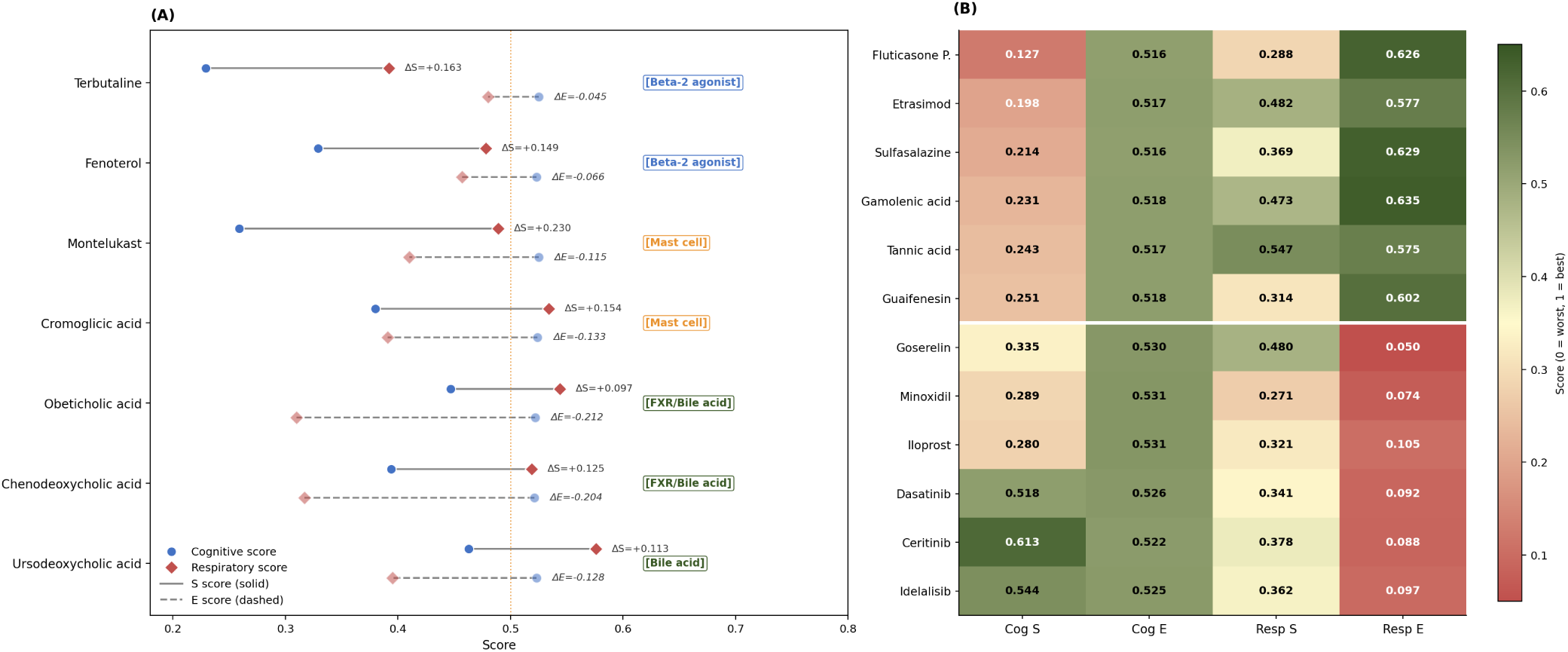
Cross-cohort score shifts and bidirectional conflict drugs. (A) Dumbbell plot for seven Long COVID-relevant cross-phenotype candidates: closed circles, cognitive scores; diamonds, respiratory scores; solid segments, S shifts; dashed segments, E shifts. All seven show positive ΔS and negative ΔE from cognitive to respiratory context. Drug class labels indicate mechanism. (B) Heatmap of bidirectional conflict drugs (avoid in one cohort, shortlist in the other). Upper block: low cognitive S but high respiratory E; lower block: high cognitive S but low respiratory E. Color scale: red=0 (worst), green=1 (best).

This dependence on phenotypes has a direct practical implication: avoid lists generated for one Long COVID presentation, as they cannot be assumed to apply to another, and the framework requires independent evaluation for each distinct patient context.

### Candidate Drugs for Long COVID

Although the primary goal of SPLIT is to identify drugs to avoid in Long COVID, the same framework naturally produces candidates with the most favorable predicted profiles. After ranking all 1,639 candidates by composite score, we applied a structured filtering step to narrow the shortlists to drugs that are realistically usable in Long COVID patients (Methods, Section). We automatically excluded agents developed exclusively for cancer treatment, drugs no longer on the market, and interventions designed only for acute or in-hospital use. We flagged but did not automatically exclude drugs that require intensive clinical monitoring or carry elevated risks in organ systems commonly affected by Long COVID, such as the heart, lungs, or nervous system; these were kept for manual review. Drugs with mechanisms directly relevant to Long COVID symptoms, including agents targeting cardiovascular dysfunction, respiratory inflammation, cognitive impairment, metabolic disturbance, or autonomic instability, were retained throughout.

After applying these filters to the top 10% of the candidates, we identified 44 Long COVID-relevant drugs on the cognitive shortlist and 62 on the respiratory shortlist (Figure 7). The 16 drugs appearing on both shortlists represent candidates with favorable profiles that are not limited to a single phenotype and, therefore, may be of broader interest in the Long COVID presentations (Figure 7, orange stars).

**Figure 7:**
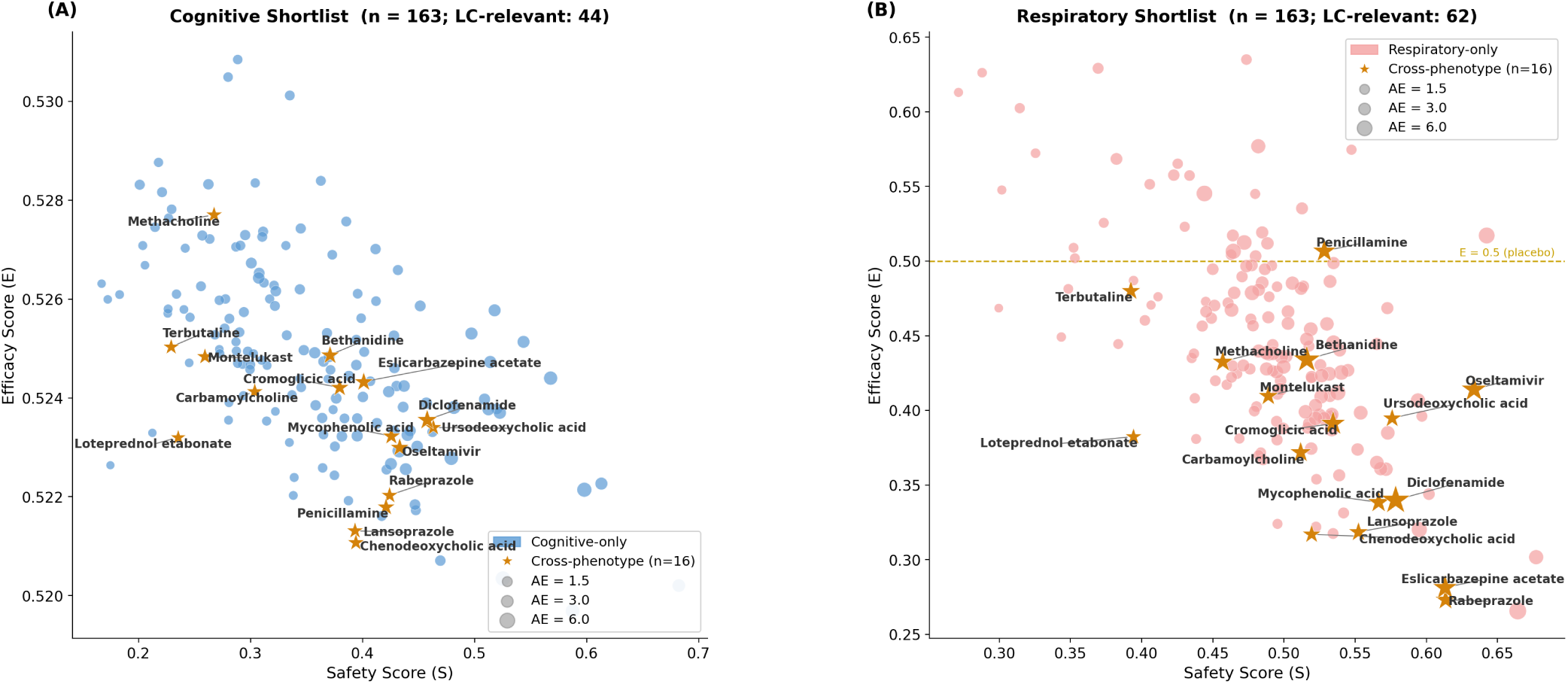
Shortlist bubble charts for cognitive and respiratory cohorts. Bubble area proportional to AE burden. Orange stars: cross-phenotype candidates (on both shortlists, n=16); cohort-specific candidates shown in cohort color. (A) Cognitive shortlist (total q10 shortlist: 163 drugs; Long COVID-relevant: 44). E is compressed (0.520–0.533); candidate separation is S-driven. (B) Respiratory shortlist (total: 163; Long COVID-relevant: 62). E ranges 0.009 to 0.635; the most favorable candidates combine S ≥ 0.45 with E *>* 0.5 (above the orange placebo line). Selected drugs labeled by name.

In the cognitive cohort, shortlisted candidates are differentiated almost entirely on the S axis, consistent with the minimal E discrimination observed across the entire panel. Drugs of particular mechanistic interest include lung vasodilators with potential vascular relevance in Long COVID (iloprost, ursodeoxycholic acid) and vasoactive agents (minoxidil). In the respiratory cohort, the shortlisted candidates cover a wide range of E (0.009–0.635), and the most favorable drugs combine moderate-to-high S with E above placebo (E *>* 0.5). Gamolenic acid and sulfasalazine reached the highest E values (0.635 and 0.629, respectively), while tannic acid combined a high E (0.575) with the highest S on the shortlist. Mast-cell stabilizers (montelukast, cromoglicic acid) and bile acid signaling modulators (ursodeoxycholic acid, chenodeoxycholic acid) appeared on both shortlists, suggesting that these candidates have favorable profiles that are consistent across phenotypic contexts. Complete shortlist tables with pharmacological annotations are provided in ST S10, ST S11, ST S18, and ST S19.

To assess whether the deprioritization predictions are consistent with established pharmacology and current clinical practice, we performed an enrichment analysis that integrates three complementary lines of evidence: statistical validation against known drug safety labels, alignment with documented pharmacological risk profiles, and cross-referencing with active Long COVID clinical programs. Complete enrichment tables are provided in the Supplementary Results (ST S18).

The alignment between computational predictions and known pharmacology was consistent across both cohorts. In the cognitive cohort, JAK inhibitors (upadacitinib, filgotinib, tofacitinib) carry FDA black box warnings for serious infections, malignancies, and cardiovascular events^41^, which is consistent with their deprioritization for predicted S grounds. Antimicrobials on the avoid list (secnidazole, ethionamide, fidaxomicin) have documented hepatotoxicity signals^42^. Oncology kinase inhibitors that flagged multiple thresholds (cobimetinib, regorafenib, sunitinib) are associated with liver toxicity, dermatological toxicity, and cardiac dysfunction^43,44^, consistent with their predicted increased burden of AE. In the respiratory cohort, deprioritized cancer agents (pazopanib, azacitidine, carboplatin, paclitaxel) carry warnings about bone marrow suppression, kidney toxicity, and nerve damage^45–47^, which aligns with their predicted near-zero E for respiratory recovery. Donepezil, deprioritized in the respiratory cohort (S=0.279), has a profile of cholinergic adverse events that may be particularly problematic in patients with autonomic instability^48^.

Together, the agreement between computational predictions, established pharmacological evidence, and the current status of the 18 flagged drugs confirms that SPLIT can serve as an up-stream candidate-evaluation tool. These results show that systematic safety-first screening can identify candidates with unfavorable predicted profiles before they use limited trial resources, and that this approach should be incorporated into the candidate selection process for Long COVID and other chronic post-infectious conditions where patient tolerability is a primary concern.

## DISCUSSION

Despite extensive computational screening since the outbreak of the pandemic^49,50^, there are no approved treatments for Long COVID^51,52^. The translation of computational hits into clinical benefit has been limited^53–55^, and we argue that the main reason is that the field continues to treat safety as a downstream filter rather than as an upstream design principle. For acute or life-threatening conditions, this ordering may be acceptable. Long COVID presents a fundamentally different challenge: patients manage chronic symptoms across multiple organ systems, often with complex medication regimens, and have limited tolerance to additional drug-related harm^1,4,11,12^. SPLIT addresses this gap by prioritizing safety in candidate selection.

However, the value of this candidate selection also depends on whether the underlying gene targets are causal drivers of disease. Most repurposed workflows are based on association signals, which can nominate targets that are downstream consequences of disease rather than modifiable drivers. Trials built on such targets fail not because the drug is ineffective, but because the target is not relevant to the disease process. Our integration of three complementary causal inference approaches addresses this problem: genetic identification via MR^20^, regulatory importance at the network level via CT^21^, and condition-specific pathway changes via DCE^22^. Each method captures a different aspect of causality, and genes supported by more than one method, including immune regulators (*STAT3*, *HIF1A*), cellular stress sensors (*TP53*, *MYC*), and the *POMC* signaling node, represent the causal drivers of the highest confidence and should be prioritized in mechanistic follow-up studies. The need to integrate all three methods is not only conceptually motivated, but also empirically necessary: at the screening threshold of *k* ≥ 11, panels built from MR or DCE genes alone produce zero drugs, while CT alone produces 991 candidates, well below the 1,639 obtained from the union of all three methods.

However, even with causally grounded targets, more is not necessarily better. Our results show that a greater interaction with the causal network signals a greater physiological disruption and, correspondingly, a higher predicted AE burden. This finding reframes the oncology kinase inhibitors that dominate target coverage rankings (often 150 to 200+ driver genes): their high mechanistic relevance is offset by toxicity profiles (hepatotoxicity, myelosuppression, cardiotoxicity)^56^ that are not compatible with long-term use in chronically symptomatic populations. The systematic exclusion of these agents demonstrates that the framework works as intended, preventing predictably harmful candidates from advancing regardless of how well they match the disease’s biology. Importantly, this trade-off between safety and coverage (*r* = −0.99) also explains why raising the gene coverage threshold does not improve the safety profile of the retained panel: higher thresholds preferentially retain the broadest-acting kinase inhibitors, which are also the least safe drugs, leaving the safety distribution unchanged from *k* = 11 to *k* = 50.

The framework also shows that deprioritization is not universal, but phenotype-specific. In the cognitive cohort, efficacy showed almost no variation between candidates, leaving safety as the only driver of deprioritization. In the respiratory cohort, efficacy varied widely, and deprioritization was primarily driven by predicted performance worse than placebo. This difference reflects genuine biological differences in how drugs interact with different patient populations and disease presentations, not a limitation of the framework. The low overlap between cohort avoid lists (11% of shared deprioritized drugs) has a clear practical consequence: deprioritization decisions generated for one Long COVID phenotype cannot be assumed to apply to another. Each clinical context requires independent evaluation. The 18 drugs deprioritized in both cohorts further illustrate this: all showed improved safety but reduced efficacy when moving from the cognitive to the respiratory context, suggesting that their unsuitability is an intrinsic property of the drugs rather than an artifact of the cohort in which they were evaluated.

The shortlist analysis reinforces this dependence on phenotypes. After structured filtering, we identified 44 candidates relevant for Long COVID on the cognitive shortlist and 62 on the respiratory shortlist. Among the 16 drugs appearing on both lists, mast cell stabilizers (montelukast, cromoglicic acid) and bile acid signaling modulators (ursodeoxycholic acid, chenodeoxycholic acid) showed consistently favorable profiles across both phenotypic contexts, suggesting that their mechanisms may address shared pathophysiological processes in Long COVID. In contrast, the 26 bidirectional conflict drugs, candidates that appear on the avoid list in one cohort but on the shortlist in the other, illustrate the practical cost of phenotype-agnostic repurposing. For example, fluticasone propionate is deprioritized due to low safety in the cognitive cohort, but appears on the respiratory shortlist due to its high efficacy, and the reverse pattern holds for cognitive-active oncology agents such as dasatinib. These conflicts would be invisible to any framework that does not evaluate candidates within phenotype-specific cohort contexts.

These computational predictions are supported by established pharmacology. Deprioritized JAK inhibitors carry FDA boxed warnings for serious infections, malignancies, cardiovascular events, and thrombosis^57–59^. Antimicrobials on the avoid list include agents with documented hepatotoxicity^42^. Cancer agents have well-known toxicities, including myelosuppression, nephrotoxicity, and neurotoxicity^45–47^. The predicted AE profiles also captured drug class-specific signatures, such as serotonergic gastrointestinal effects for vortioxetine and CYP3A4-mediated interactions for ritonavir, suggesting that the model captures genuine toxicity patterns rather than statistical artifacts.

Perhaps more importantly, some drugs on our list of avoidance are currently under active investigation for Long COVID. The JAK inhibitors baricitinib and upadacitinib have advanced to phase 3 despite predicted unfavorable profiles, and FDA boxed warnings^41,57,58,60,61^. Metformin, ivermectin, and ritonavir, tested in major trial programs, also appear on the avoid lists^25,62–64^. This does not mean that these trials are misguided; mechanistic rationale may justify investigating higher-risk agents in selected contexts, and our predictions require prospective validation. However, trial designs should include improved safety monitoring for computationally flagged agents or provide clear justification for the predicted risks given the anticipated benefit.

As with any *in silico* screening approach, our predictions depend on the training knowledge graph and do not model dose-response relationships, schedule variations, or adherence effects; these can only be resolved through prospective clinical testing. The causal driver gene set reflects current Long COVID multi-omics datasets and will benefit from updates as larger cohorts become available. Within these boundaries, SPLIT is designed as a decision support tool for early candidate screening, not as a replacement for clinical judgment or regulatory evaluation. Its value lies in preventing predictably problematic candidates from using limited trial resources, and in focusing effort on candidates with more favorable predicted profiles.

Together, these findings position SPLIT as a practical and clinically relevant framework for identifying safer and more context-appropriate repurposing candidates in Long COVID.

## Data Availability

All analyses were performed using reproducible workflows in Python 3.11, with environment specifications captured through conda. Data processing, network construction, MR pipelines, DCE implementation, CT analyses, drug--gene mapping, and PlaNet inference are encapsulated in modular scripts and notebooks executable on standard compute or high-performance clusters. All source code, configuration files, and minimal processed input required to reproduce the results are publicly available at \url{https://github.com/SindyPin/SPLIT}, following the principles of open science and FAIR.

https://github.com/SindyPin/SPLIT

## RESOURCES

### Lead Contact

Thuc Duy Le.

### Materials Availability

This study did not generate new unique reagents.

### Data and Code Availability

All analyses were performed using reproducible workflows in Python 3.11, with environment specifications captured through conda. Data processing, network construction, MR pipelines, DCE implementation, CT analyses, drug–gene mapping, and PlaNet inference are encapsulated in modular scripts and notebooks executable on standard compute or high-performance clusters. All source code, configuration files, and minimal processed input required to reproduce the results are publicly available at https://github.com/SindyPin/SPLIT, following the principles of open science and FAIR.

## ACKNOWLEDGMENTS

This work was partly supported by the Australian Research Council Discovery Project under Grant DP230101122 and the University Presidents Scholarship (UPS) stipend.

## AUTHOR CONTRIBUTIONS

**Conceptualization:** Thuc Duy Le. **Data Curation:** Sindy Pinero. **Formal Analysis:** Sindy Pinero. **Funding Acquisition:** Thuc Duy Le. **Investigation:** Sindy Pinero. **Methodology:** Sindy Pinero and Thuc Duy Le. **Project Administration:** Thuc Duy Le. **Resources:** Lin Liu, Jiuyong Li, Sang Hong Lee, and Thuc Duy Le. **Code:** Sindy Pinero. **Supervision:** Xiaomei Li, Lin Liu, Jiuyong Li, Sang Hong Lee, and Thuc Duy Le. **Validation:** Lin Liu, Sang Hong Lee, and Thuc Duy Le. **Visualization:** Sindy Pinero. **Writing and Original Draft:** Sindy Pinero. **Writing and Review & Editing:** Sindy Pinero, Xiaomei Li, Lin Liu, Jiuyong Li, Sang Hong Lee, and Thuc Duy Le.

## DECLARATION OF GENERATIVE AI AND AI-ASSISTED TECH-NOLOGIES

During the preparation of this work, the author(s) used ChatGPT (https://chat.openai.com) and Claude (https://claude.ai) to improve the writing and clarity of the manuscript. After using this tool or service, the author(s) reviewed and edited the content as needed and take(s) full responsibility for the content of the publication.

## SUPPLEMENTAL INFORMATION INDEX

To support reproducibility and allow for subsequent inspection, we provide the complete set of intermediate and final outputs in the Supplementary Materials. Table summarizes the organization of the supplementary tables and data files.

| Item | Description |
| --- | --- |
| **Supplementary Methods and Results document** |  |
| SM | Full supplementary methods (Sections 1–8: causal gene discovery, clinical trial parsing, PlaNet architecture, AE burden scoring, composite ranking, avoid list definitions, filtering rules, pharmacology enrichment) and supplementary results (Sections 9–18: glossary, gene evidence tables, mapping details, counterfactual predictions, AE signatures, avoid list concordance, dimensional risk profiles, secondary screening, shared deprioritized drugs, score distributions) |
| **Shared resources** |  |
| ST S1 | Full causal gene list with evidence labels from CT/MR/DCE (1,725 genes) |
| ST S2 | Complete drug to causal gene mapping table across DrugBank, ChEMBL, TTD, DrugCentral, and GtoPdb |
| ST S3 | Ground truth and AE probability tables for counterfactual scenarios (Niagen, Ritonavir, Vortioxetine, Leronlimab) |
| **NCT04809974 (Cognitive cohort)** |  |
| ST S4 | Raw clinical trial data retrieved from ClinicalTrials.gov |
| ST S5 | Parsed trial representations and cohort descriptors |
| ST S6 | PlaNet predictions: S/E/AE scores for all screened candidates |
| ST S7 | Mapped AE outputs: code/label tables and summaries |
| ST S8 | Full composite ranking table with normalized S/E/AE scores |
| ST S9 | Avoid lists: primary composite, threshold, delta, and sensitivity analyses |
| ST S10 | Enriched pharmacology table with targets, potency metrics, and translational annotations |
| ST S11 | Master tables: consolidated outputs for the cognitive cohort |
| **NCT04880161 (Respiratory cohort)** |  |
| ST S12 | Raw clinical trial data retrieved from ClinicalTrials.gov |
| ST S13 | Parsed trial representations and cohort descriptors |
| ST S14 | PlaNet predictions: S/E/AE scores for all screened candidates |
| ST S15 | Mapped AE outputs: code/label tables and summaries |
| ST S16 | Full composite ranking table with normalized S/E/AE scores |
| ST S17 | Avoid lists: primary composite, threshold, delta, and sensitivity analyses |
| ST S18 | Enriched pharmacology table with targets, potency metrics, and translational annotations |
| ST S19 | Master tables: consolidated outputs for the respiratory cohort |
| **Literature and clinical context** |  |
| ST S20 | Long COVID therapeutic context references and cross referencing of avoid list drugs against active clinical trials and preclinical programs |
| ST S21 | More Details for Methods and Results |

---

| Item | Description |
| --- | --- |
| **Supplementary Methods and Results document** |  |
| SM | Full supplementary methods (Sections 1–8: causal gene discovery, clinical trial parsing, PlaNet architecture, AE burden scoring, composite ranking, avoid list definitions, filtering rules, pharmacology enrichment) and supplementary results (Sections 9–18: glossary, gene evidence tables, mapping details, counterfactual predictions, AE signatures, avoid list concordance, dimensional risk profiles, secondary screening, shared deprioritized drugs, score distributions) |
| **Shared resources** |  |
| ST S1 | Full causal gene list with evidence labels from CT/MR/DCE (1,725 genes) |
| ST S2 | Complete drug to causal gene mapping table across DrugBank, ChEMBL, TTD, DrugCentral, and GtoPdb |
| ST S3 | Ground truth and AE probability tables for counterfactual scenarios (Niagen, Ritonavir, Vortioxetine, Leronlimab) |
| **NCT04809974 (Cognitive cohort)** |  |
| ST S4 | Raw clinical trial data retrieved from ClinicalTrials.gov |
| ST S5 | Parsed trial representations and cohort descriptors |
| ST S6 | PlaNet predictions: S/E/AE scores for all screened candidates |
| ST S7 | Mapped AE outputs: code/label tables and summaries |
| ST S8 | Full composite ranking table with normalized S/E/AE scores |
| ST S9 | Avoid lists: primary composite, threshold, delta, and sensitivity analyses |
| ST S10 | Enriched pharmacology table with targets, potency metrics, and translational annotations |
| ST S11 | Master tables: consolidated outputs for the cognitive cohort |
| **NCT04880161 (Respiratory cohort)** |  |
| ST S12 | Raw clinical trial data retrieved from ClinicalTrials.gov |
| ST S13 | Parsed trial representations and cohort descriptors |
| ST S14 | PlaNet predictions: S/E/AE scores for all screened candidates |
| ST S15 | Mapped AE outputs: code/label tables and summaries |
| ST S16 | Full composite ranking table with normalized S/E/AE scores |
| ST S17 | Avoid lists: primary composite, threshold, delta, and sensitivity analyses |
| ST S18 | Enriched pharmacology table with targets, potency metrics, and translational annotations |
| ST S19 | Master tables: consolidated outputs for the respiratory cohort |
| **Literature and clinical context** |  |
| ST S20 | Long COVID therapeutic context references and cross referencing of avoid list drugs against active clinical trials and preclinical programs |

## STAR METHODS

### Causal Gene Discovery

We identified likely Long COVID driver genes by combining three complementary causal inference strategies, each designed to capture a different aspect of driver biology.

TWMR identifies genes whose genetically predicted expression has a causal effect on Long COVID risk, providing a human genetic basis for target selection ^20,65^. CT identifies genes that are essential for maintaining network function: specifically, genes whose removal increases the control input needed to steer the system, reflecting their regulatory importance within the protein interaction network ^21,65^. DCE measures how gene-to-gene regulatory relationships within pathways change in Long COVID patients compared to healthy controls, revealing condition-specific rewiring of the pathway structure ^22,66^.

All analyses used harmonized identifiers, consistent quality control, and uniform multiple testing correction. We retained genes supported by at least one method and not contradicted by sensitivity checks. The complete annotated gene list is provided in ST S1; detailed method descriptions are in SM, Sections 1.1–1.4.

### Drug to Gene Mapping

We linked each driver gene to the drugs that act on it using five curated pharmacological databases: DrugBank ^67^, ChEMBL ^68^, the Therapeutic Target Database ^69^, DrugCentral ^70^, and the Guide to PHARMACOLOGY ^71^. For each driver gene, we retrieved approved drugs and advanced investigational agents with direct target annotations, including agonists, antagonists, inhibitors, antibodies, and modulators. Drug and gene identifiers were standardized across databases, and duplicate or obsolete entries were removed. This step ensures that every drug on the screening panel has a documented mechanistic link to at least one causal Long COVID driver gene. The complete mapping table is provided in ST S2.

### Drug Panel Threshold Justification

After mapping, 19,172 unique drugs had at least one causal gene target. To focus on drugs with meaningful mechanistic coverage, we applied a minimum target threshold of *k* ≥ 11 causal genes, reducing the panel to 1,776 candidates. We justified this threshold through four lines of evidence.

First, we performed a hypergeometric enrichment test for each of the 19,172 drugs, asking whether their overlap with the 1,725 Long COVID causal genes is greater than expected by chance against a background of 20,000 human genes ^72^. Multiple testing corrections were applied using the Benjamini-Hochberg procedure. This test provides a statistically grounded basis for the threshold, independent of the downstream scoring step.

Second, we examine the distribution of causal gene targets across the entire set of drugs to assess where *k* = 11 falls within that distribution and to track how the panel size changes as the threshold increases. This confirmed that the results are robust to moderate variation around the chosen value.

Third, we tested whether raising the threshold improves the safety profile of the retained panel by comparing the safety score distributions at thresholds from *k* = 11 to *k* = 50 in both cohorts.

Finally, we assessed the contribution of each causal inference method to the panel by comparing the panel sizes obtained from CT, MR, and DCE gene sets individually against the union of all three. This determined whether integrating multiple methods is necessary to achieve a usable screening panel at this threshold.

### Clinical Trial Processing and PlaNet Prediction

We used PlaNet ^23^, a geometric deep learning model trained on a large-scale clinical knowledge graph, to predict adverse event profiles, global safety scores, and comparative efficacy for each candidate drug.

PlaNet represents each clinical trial arm as a node in a knowledge graph. Each node is connected by typed edges to drugs, diseases, outcomes, and population descriptors that define the trial protocol. The model then propagates information across this graph and across biomedical background networks, including drug similarity hierarchies, protein interaction networks, and gene function ontologies, using a relational graph convolutional encoder. It was pretrained using a self-supervised link-prediction objective and then fine-tuned on adverse-event enrichment and survival outcome tasks derived from completed clinical trials.

The clinical trial records for both cohorts were retrieved from ClinicalTrials.gov and parsed into structured representations compatible with PlaNet, following and extending the processing pipeline described by the original authors ^23^. We applied the pretrained model without further retraining. Full details of the parsing pipeline, knowledge graph projection, and model architecture are provided in SM, Sections 2–3. Raw trial data and parsed representations are in ST S4 and ST S5 (NCT04809974) and ST S12 and ST S13 (NCT04880161); prediction outputs are in ST S6 and ST S14.

### Counterfactual Substitution

To evaluate each candidate drug under matched conditions, we used counterfactual substitution. In this design, the experimental drug in a real clinical trial record is replaced with the candidate drug, while everything else about the trial context remains the same: disease indication, eligibility criteria, enrollment, demographics, and comparator structure. This ensures that any differences in predicted safety, efficacy, and adverse-event burden reflect the drug itself rather than cohort or protocol variation.

To prevent real trial outcomes from influencing predictions, we removed all field values derived from results from the trial record and replaced outcome summaries with neutral placeholder statements before generating counterfactual predictions. Details of the replacement procedure and leakage controls are provided in SM, Section 2.7.

### Evaluation Design

Our evaluation uses a counterfactual design based on real clinical trial cohorts. For a fixed cohort and protocol, we compared the predicted S, AE, and E for the original trial intervention with those obtained after substituting each candidate drug in the same context (Section).

We used a template and sensitivity design to test whether the predictions depend on the specific cohort. The template cohort served as the primary evaluation context, while the sensitivity cohort, with distinct phenotypic characteristics, tested whether deprioritization patterns generalize across Long COVID presentations.

NCT04809974 (Niagen; cognitive and brain fog phenotype; *N* = 72; mean age 46 years; 70% female; 91% White) served as the template cohort. NCT04880161 (Ampion; respiratory phenotype; *N* = 32; mean age 52 years; 56% female; 75% White, 25% Black) served as the sensitivity cohort. Table 5 summarizes the characteristics of each cohort; raw clinical trial data are provided in ST S4 (NCT04809974) and ST S12 (NCT04880161), with parsed cohort representations in ST S5 and ST S13, respectively.

**Table 5:** Characteristics of Long COVID clinical trial cohorts used for template evaluation and sensitivity analysis.

| Characteristic | NCT04809974 (Template) | NCT04880161 (Sensitivity) |
| --- | --- | --- |
| **Intervention** |  |  |
| Drug | Niagen (Nicotinamide Riboside) | Ampion (Inhaled Biologic) |
| Primary target domain | Cognitive (brain fog) | Respiratory |
| **Population** |  |  |
| Sample size ($N$) | 72 | 32 |
| Age range | 18–65 | 18+ |
| Age, mean (SD) | 45.9 (13.4) | 52.1 (13.0) |
| Female, $N$ (%) | 42 (72.4) | 18 (56.3) |
| BMI, mean (SD) | N/A | 33.4 (7.1) |
| **Race/ethnicity** |  |  |
| White, $N$ (%) | 53 (91) | 24 (75) |
| Black, $N$ (%) | 2 (3) | 8 (25) |
| Hispanic, $N$ (%) | 5 (9) | NR |
| **Eligibility criteria** |  |  |
| Time since COVID | $\geq 2$ months | $\geq 4$ weeks |
| Baseline symptoms | Brain fog + $\geq 2$ neuro/physical symptoms | Respiratory $\geq 4$ weeks; $\geq 2$ symptoms score $\geq 2$ |
| Key exclusions | CNS disease, major psychiatric illness | Severe COPD, CKD, liver disease, CFS |

**Abbreviations:** SD, standard deviation; NR, not reported; CNS, central nervous system; COPD, chronic obstructive pulmonary disease; CKD, chronic kidney disease; CFS, chronic fatigue syndrome; BMI, body mass index.

For counterfactual evaluation, we selected three candidate drugs that cover a range of mechanistic targets: Leronlimab (1 causal gene target), Vortioxetine (11 targets), and Ritonavir (70 targets) ^73^. Each was substituted into the template cohort to generate matched predictions. Throughout, the emphasis is on avoidance rather than discovery: drugs with a lower predicted S or higher AE burden relative to the original trial intervention were flagged for deprioritization.

### Safety, Adverse Event, and Efficacy Scoring

For each candidate drug, PlaNet ^23^ produces three outputs, all defined relative to a placebo baseline within the same cohort context: a global safety score (S), a comparative efficacy score (E), and the probability of enrichment for each adverse event category. The full definitions of these outputs are provided in SM, Section 3.

To summarize the predicted toxicity in a way that gives greater weight to clinically severe and Long COVID-relevant events, we calculated a weighted adverse event burden score (AE). This score combines PlaNet’s per-category probabilities with severity weights aligned to the CTCAE grading system ^74^ and Long COVID relevance weights. We then combined S, E, and the inverse AE burden into a single composite ranking score *C*, with default weights *w_S_* = 0.4, *w_E_*= 0.4, and *w_AE_* = 0.2. These weights reflect the safety-first principle for SPLIT while treating efficacy as a co-dominant signal. When comparative efficacy was unavailable for a given cohort, the weights were renormalized over the remaining components. Score definitions, weighting schemes, and normalization procedures are detailed in SM, Sections 4–5; predicted scores and mapped adverse event output are in ST S6 and ST S7 (NCT04809974) and ST S14 and ST S15 (NCT04880161); full ranking tables are in ST S8 and ST S16.

### Deprioritization Avoid Lists

We generated three complementary avoid lists, each designed to capture a different aspect of unfavorable predicted profiles.

The **primary composite avoid list** ranks all screened drugs by composite score *C* and flags the lowest-scoring candidates at three levels of stringency: the worst 5% (q05), the worst 10% (q10, used as default threshold), and the worst 20% (q20).

The **secondary threshold list** provides an independent check by flagging drugs with extreme values in individual metrics at q20 thresholds for the AE burden, S, or E. The number of triggered flags per drug ranges from 1 to 3 (*n*_extremes_ ∈ {1, 2, 3}), with higher counts indicating stronger evidence of an unfavorable profile.

The **counterfactual delta list** identifies drugs that perform worse than the original trial intervention after substitution on any individual metric (Δ*S <* 0, Δ*E <* 0, or Δ*AE >* 0), with candidates in the worst decile flagged for deprioritization. The number of worsened metrics per drug (*n*_delta_ ∈ {1, 2, 3}) is tracked to identify candidates with broadly inferior profiles.

Drugs appearing on multiple lists represent higher-confidence deprioritization targets. Sensitivity across stringency levels was assessed by repeating all analyses at q05, q10, and q20; candidates that remain flagged at all levels form a conservative avoid core. Complete lists and concordance metrics are provided in ST S9 and ST S17; detailed definitions are provided in SM, Section 6.

### Filtering for Long COVID Candidates

After scoring, we applied a structured post-processing step to generate candidate shortlists suited to Long COVID. We first restricted the pool to top-performing candidates by cohort rank, then optionally applied thresholds on normalized score components and a Pareto filter to retain candidates that are not outperformed on all metrics simultaneously by another drug.

Within this filtered set, we applied three rule categories. **Hard-block rules** automatically excluded agents that are not appropriate for Long COVID management, including oncology-specific agents without established non-oncology indications, acute-only interventions unsuitable for chronic outpatient use, and withdrawn drugs. **Soft-block rules** flagged agents that require intensive monitoring or carry elevated risks in organ systems commonly affected by Long COVID for manual review, without automatic exclusion. Keep-hint rules preserved visibility for candidates with mechanisms directly relevant to Long COVID symptoms, including cardiopulmonary, respiratory, neurocognitive, metabolic, autonomic, and supportive categories. Detailed rule definitions are provided in SM, Section 7.

### Pharmacology Enrichment

To place the computational predictions in the context of established pharmacological evidence, we enriched each candidate drug with information retrieved programmatically from PubChem ^75,76^, ChEMBL ^77^, Open Targets ^78^, and DailyMed ^79^. For each drug, we collected the approval status and clinical phase, the physicochemical properties (molecular weight, lipophilicity, polar surface area, hydrogen bond donors and acceptors), the safety label signals including FDA boxed warnings, the curated potency records (IC_50_, EC_50_, *K_i_*, *K_d_*), and the mechanism of action annotations. As a development screening, we calculated the Lipinski Rule of five violations ^80^. Complete enrichment tables are provided in ST S10 and ST S18; consolidated outputs for each cohort are in ST S11 and ST S19. The enrichment protocol is described in SM, Section 8.

## References

1. Davis, H.E., McCorkell, L., Vogel, J.M., and Topol, E.J. (2023). Long covid: major findings, mechanisms and recommendations. Nature Reviews Microbiology 21, 133–146. URL: https://www.nature.com/articles/s41579-023-00884-5. doi: 10.1038/s41579-023-00884-5.

2. Lopez-Leon, S., Wegman-Ostrosky, T., Perelman, C., and, et al. (2021). More than 50 long-term effects of covid-19: a systematic review and meta-analysis. Scientific Reports 11, 16144. URL: https://www.nature.com/articles/s41598-021-95565-8. doi: 10.1038/s41598-021-95565-8.

3. Soriano, J.B., Murthy, S., Marshall, J.C., Relan, P., and Diaz, J.V. (2022). A clinical case definition of post covid-19 condition by a delphi consensus. The Lancet Infectious Diseases 22, e102–e107. URL: https://www.thelancet.com/journals/laninf/article/PIIS1473-3099(21)00703-9/fulltext. doi: 10.1016/S1473-3099(21)00703-9.

4. World Health Organization (2025). Post covid-19 condition (long covid). Fact sheet. URL: https://www.who.int/news-room/fact-sheets/detail/post-covid-19-condition-(long-covid).

5. US Food and Drug Administration. Long covid: Voice of the patient report. Tech. Rep. US Food and Drug Administration (2023). URL: https://www.fda.gov/media/177092/download.

6. Zeraatkar, D., Ling, M., Kirsh, S., Jassal, T., Shahab, M., Walch, A., Talukdar, J.R. et al. (2024). Interventions for the management of long covid (post-covid condition): living systematic review. BMJ 387, e081318. URL: https://www.bmj.com/content/387/bmj-2024-081318. doi: 10.1136/bmj-2024-081318.

7. Ivlev, I., Wagner, J., Phillips, T., and Treadwell, J.R. (2025). Interventions for long covid: A narrative review. Journal of General Internal Medicine 40, 2005–2023. URL: https://link.springer.com/article/10.1007/s11606-024-09254-z. doi: 10.1007/s11606-024-09254-z.

8. Tan, C., Meng, J., Dai, X., He, B., Liu, P., Wu, Y., Xiong, Y., Yin, H., Wang, S., and Gao, S. (2025). Effects of therapeutic interventions on long covid: a meta-analysis of randomized controlled trials. EClinicalMedicine 87, 103412. URL: 10.1016/j.eclinm.2025.103412. doi: 10.1016/j.eclinm.2025.103412.

9. Centers for Disease Control and Prevention (2025). Long covid clinical guidance. URL: https://www.cdc.gov/long-covid/hcp/clinical-guidance/index.html updated 24 July 2025; Accessed 27 Jan 2026.

10. National Institute for Health and Care Excellence (2024). Covid-19 rapid guideline: managing the long-term effects of covid-19 (ng188). URL: https://www.nice.org.uk/guidance/ng188 last updated 25 Jan 2024; Accessed 27 Jan 2026.

11. Michael, H.U., Brouillette, M.J., Fellows, L.K., and Mayo, N.E. (2024). Medication utilization patterns in patients with post-covid syndrome (pcs): Implications for polypharmacy and drug–drug interactions. Journal of the American Pharmacists Association 64, 102083. URL: https://www.sciencedirect.com/science/article/pii/S1544319124001031. doi: 10.1016/j.japh.2024.102083.

12. Chen, Z., Tian, F., and Zeng, Y. (2023). Polypharmacy, potentially inappropriate medications, and drug–drug interactions in older covid-19 inpatients. BMC Geriatrics 23, 774. URL: https://bmcgeriatr.biomedcentral.com/articles/10.1186/s12877-023-04487-9. doi: 10.1186/s12877-023-04487-9.

13. Ashburn, T.T., and Thor, K.B. (2004). Drug repositioning: identifying and developing new uses for existing drugs. Nature Reviews Drug Discovery 3, 673–683. URL: https://www.nature.com/articles/nrd1468. doi: 10.1038/nrd1468.

14. Pushpakom, S., Iorio, F., Eyers, P.A., Escott, K.J., Hopper, S., Wells, A., Doig, A., Guilliams, T., Latimer, J., McNamee, C. et al. (2019). Drug repurposing: progress, challenges and recommendations. Nature Reviews Drug Discovery 18, 41–58. URL: https://www.nature.com/articles/nrd.2018.168. doi: 10.1038/nrd.2018.168.

15. Pinzi, L., Bisi, N., and Rastelli, G. (2024). How drug repurposing can advance drug discovery: challenges and opportunities. Frontiers in Drug Discovery 4, 1460100. URL: 10.3389/fddsv.2024.1460100. doi: 10.3389/fddsv.2024.1460100.

16. Recino, A., Rayner, M.L.D., Rohn, J.L., Della Pasqua, O., and UCL Repurposing TIN Committee (2025). Therapeutic innovation in drug repurposing: challenges and opportunities. Drug Discovery Today 30, 104390. URL: 10.1016/j.drudis.2025.104390. doi: 10.1016/j.drudis.2025.104390.

17. Morselli Gysi, D., do Valle, Í., Zitnik, M., Ameli, A., Gan, X., Varol, O., Ghiassian, S.D., Patten, J.J., Davey, R.A., Loscalzo, J., and Barabási, A.L. (2021). Network medicine framework for identifying drug-repurposing opportunities for covid-19. Proceedings of the National Academy of Sciences of the United States of America 118, e2025581118. doi: 10.1073/pnas.2025581118.

18. Zhou, Y., Hou, Y., Shen, J., Huang, Y., Martin, W., and Cheng, F. (2020). Network-based drug repurposing for novel coronavirus 2019-ncov/sars-cov-2. Cell Discovery 6, 14. URL: https://www.nature.com/articles/s41421-020-0153-3. doi: 10.1038/s41421-020-0153-3.

19. Zhang, R., Hristovski, D., Schutte, D., Kastrin, A., Fiszman, M., and Kilicoglu, H. (2021). Drug repurposing for covid-19 via knowledge graph completion. Journal of Biomedical Informatics 115, 103698. doi: 10.1016/j.jbi.2021.103698.

20. Gleason, K.J., Yang, F., and Chen, L.S. (2021). A robust two-sample transcriptome-wide Mendelian Randomization method integrating GWAS with multi-tissue eQTL summary statistics. Genetic Epidemiology 45, 353–371. doi: 10.1002/gepi.22380.

21. Vinayagam, A., Gibson, T.E., Lee, H.J., Yilmazel, B., Roesel, C., Hu, Y., Kwon, Y., Sharma, A., Liu, Y.Y., Perrimon, N., and Barabási, A.L. (2016). Controllability analysis of the directed human protein interaction network identifies disease genes and drug targets. Proceedings of the National Academy of Sciences of the United States of America 113, 4976–4981. doi: 10.1073/pnas.1603992113.

22. Jablonski, K.P., Pirkl, M., Ćevid, D., Bühlmann, P., and Beerenwinkel, N. (2022). Identifying cancer pathway dysregulations using differential causal effects. Bioinformatics 38, 1550–1559. doi: 10.1093/bioinformatics/btab847.

23. Brbic, M., Yasunaga, M., Agarwal, P., and Leskovec, J. (2024). Planet: Predicting population response to drugs via clinical knowledge graph. medRxiv. URL: https://www.medrxiv.org/content/10.1101/2024.01.01.000001v1. doi: 10.1101/2024.01.01.000001.

24. Sitbon, O., Channick, R., Chin, K.M., Frey, A., Gaine, S., Galie, N., Ghofrani, H.A., Hoeper, M.M., Lang, I.M., Preiss, R., Rubin, L.J., Di Scala, L., Tapber, N., Torbicki, A., and Simonneau, G. (2015). Selexipag for the treatment of pulmonary arterial hypertension. New England Journal of Medicine 373, 2522–2533. doi: 10.1056/NEJMoa1503184.

25. Bramante, C.T. et al. (2023). Outpatient treatment of covid-19 and incidence of post-covid-19 condition (long covid) in a randomised, placebo-controlled trial. The Lancet Infectious Diseases. URL: https://www.thelancet.com/journals/laninf/article/PIIS1473-3099(23)00299-2/fulltext. doi: 10.1016/S1473-3099(23)00299-2. COVID-OUT trial reports metformin/ivermectin arms and long COVID incidence outcomes.

26. Geng, L.N., Bonilla, H., Hedlin, H. et al. (2024). Nirmatrelvir-ritonavir and symptoms in adults with postacute sequelae of SARS-CoV-2 infection: the STOP-PASC randomized clinical trial. JAMA Intern Med 184, 1024–1034. doi: 10.1001/jamainternmed.2024.2007.

27. ClinicalTrials.gov (2023). Reverse-long covid-19 with baricitinib (nct05858515). URL: https://clinicaltrials.gov/study/NCT05858515 clinical trial registration; accessed 2025-12-21.

28. ClinicalTrials.gov (2025). Long covid (lc)-revitalize (nct06928272). URL: https://clinicaltrials.gov/study/NCT06928272 clinical trial registration; accessed 2025-12-21.

29. ClinicalTrials.gov (2022). Ivabradine for long-term effects of covid-19 with pots (nct05481177). URL: https://clinicaltrials.gov/study/NCT05481177 clinical trial registration; accessed 2025-12-21.

30. ClinicalTrials.gov (2024). Recover-autonomic (ivabradine) (nct06305806). URL: https://clinicaltrials.gov/study/NCT06305806 clinical trial registration; accessed 2025-12-21.

31. ClinicalTrials.gov (2024). Recover-sleep: Platform protocol (nct06404086). URL: https://clinicaltrials.gov/study/NCT06404086 clinical trial registration; accessed 2025-12-21.

32. Bonilla, H., Peluso, M.J., Rodgers, K., Aberg, J.A., Patterson, T.F. et al. (2023). Therapeutic trials for long covid-19: A call to action from the interventions taskforce of the recover initiative. Frontiers in Immunology 14, 1129459. URL: 10.3389/fimmu.2023.1129459. doi: 10.3389/fimmu.2023.1129459.

33. O’Kelly, B., Vidal, L., McHugh, T. et al. (2022). Safety and efficacy of low dose naltrexone in a long COVID cohort: an interventional pre-post study. Brain Behav Immun Health 24, 100485. doi: 10.1016/j.bbih.2022.100485.

34. Pretorius, E., Laubscher, G.J., Kell, D.B. et al. (2023). Treatment of long COVID symptoms with triple anticoagulant therapy. Research Square. doi: 10.21203/rs.3.rs-2697680/v1. Preprint.

35. Fesharaki-Zadeh, A., Lowe, N., and Arnsten, A.F.T. (2023). Clinical experience with the *α*2a-adrenoceptor agonist, guanfacine, and n-acetylcysteine for the treatment of cognitive deficits in “long-covid19”. Neuroimmunology Reports 3, 100154. doi: 10.1016/j.nerep.2022.100154.

36. ClinicalTrials.gov (2025). Impact-lc: Maraviroc and atorvastatin for long covid (nct06974084). URL: https://clinicaltrials.gov/study/NCT06974084 clinical trial registration; accessed 2025-12-21.

37. Reinfeld, S. (2023). Can bupropion treat COVID-19–induced brain fog? a case series. Int Clin Psychopharmacol 38, 189–191. doi: 10.1097/YIC.0000000000000436.

38. ClinicalTrials.gov (2024). Effect of vitamin d supplementation on glutathione peroxidase (gpx) activity, interleukin-6 (il-6) levels and clinical outcomes in post-covid-19 patients. https://clinicaltrials.gov/study/NCT06419712. Clinical trial registration, NCT06419712. Intervention includes cholecalciferol.

39. Charoenporn, V., Tungsukruthai, P., Teacharushatakit, P. et al. (2024). Effects of an 8-week high-dose vitamin d supplementation on fatigue and neuropsychiatric manifestations in post-covid syndrome: A randomized controlled trial. Psychiatry Clin Neurosci 78, 595–604. doi: 10.1111/pcn.13716.

40. Conze, D., Brenner, C., and Kruger, C.L. (2019). Safety and metabolism of long-term administration of NIAGEN (nicotinamide riboside chloride) in a randomized, double-blind, placebo-controlled clinical trial of healthy overweight adults. Scientific Reports 9, 9772. doi: 10.1038/s41598-019-46120-z.

41. U.S. Food and Drug Administration (2021). Fda requires warnings about increased risk of serious heart-related events, cancer, blood clots, and death for jak inhibitors. URL: https://www.fda.gov/drugs/drug-safety-and-availability/fda-requires-warnings-about-increased-risk-serious-heart-related-events-cancer-blood-clots-and-death drug Safety Communication; Accessed 27 Jan 2026.

42. National Institute of Diabetes and Digestive and Kidney Diseases (2020). Ethionamide. URL: https://www.ncbi.nlm.nih.gov/books/NBK548025/ liverTox: Clinical and Research Information on Drug-Induced Liver Injury; Accessed 27 Jan 2026.

43. Ghatalia, P., Je, Y., Kaymakcalan, M.D., Sonpavde, G.P., and Choueiri, T.K. (2015). Hepatotoxicity with vascular endothelial growth factor receptor tyrosine kinase inhibitors: A meta-analysis of randomized clinical trials. Critical Reviews in Oncology/Hematology 93, 257–276. doi: 10.1016/j.critrevonc.2014.11.006.

44. Abdel-Rahman, O., and Fouad, M. (2014). Risk of cardiovascular toxicities in patients with solid tumors treated with sunitinib, axitinib, cediranib or regorafenib: An updated systematic review and comparative meta-analysis. Critical Reviews in Oncology/Hematology 92, 194–207. doi: 10.1016/j.critrevonc.2014.06.003.

45. U.S. Food and Drug Administration (2012). Votrient (pazopanib) prescribing information. URL: https://www.accessdata.fda.gov/drugsatfda_docs/label/2012/022465s-010S-012lbl.pdf boxed warning: hepatotoxicity; Accessed 27 Jan 2026.

46. U.S. Food and Drug Administration (2010). Paraplatin (carboplatin) prescribing information. URL: https://www.accessdata.fda.gov/drugsatfda_docs/label/2010/020452s005lbl.pdf warnings include severe myelosuppression; nephrotoxicity risk with concomitant nephrotoxins; Accessed 27 Jan 2026.

47. U.S. Food and Drug Administration (2023). Paclitaxel protein-bound particles for injectable suspension (albumin-bound) prescribing information. URL: https://www.accessdata.fda.gov/drugsatfda_docs/label/2023/216338s000lbl.pdf warnings include severe myelosuppression; peripheral neuropathy/neurotoxicity discussed in labeling; Accessed 27 Jan 2026.

48. Park-Wyllie, L.Y., Mamdani, M.M., Li, P., Gill, S.S., Laupacis, A., and Juurlink, D.N. (2009). Cholinesterase inhibitors and hospitalization for bradycardia: A population-based study. PLoS Medicine 6, e1000157. doi: 10.1371/journal.pmed.1000157.

49. Galindez, G., Matschinske, J., Rose, T.D., Sadegh, S., Salgado-Albarrán, M., Späth, J., Baumbach, J., and Pauling, J.K. (2021). Lessons from the covid-19 pandemic for advancing computational drug repurposing strategies. Nature Computational Science 1, 33–41. URL: https://www.nature.com/articles/s43588-020-00007-6. doi: 10.1038/s43588-020-00007-6.

50. Zhou, Y., Wang, F., Tang, J., Nussinov, R., and Cheng, F. (2020). Artificial intelligence in covid-19 drug repurposing. The Lancet Digital Health 2, e667–e676. URL: https://www.thelancet.com/journals/landig/article/PIIS2589-7500(20)30192-8/fulltext. doi: 10.1016/S2589-7500(20)30192-8.

51. NPR (2024). Long COVID patients push for NIH research funds to focus on treatments. NPR Shots Health News. Available at: https://www.npr.org/sections/shots-health-news/2024/11/25/nx-s1-5199994/long-covid-patients-nih-research-treatments.

52. National Institutes of Health (2024). RECOVER: Researching COVID to enhance recovery. https://recovercovid.org.

53. Camps, I., Künzel, S.R., Schubert, M., and Singh, R.K. (2025). Editorial: Opportunities and challenges in drug repurposing. Frontiers in Pharmacology 16, 1709217. URL: https://www.frontiersin.org/journals/pharmacology/articles/10.3389/fphar.2025.1709217/full. doi: 10.3389/fphar.2025.1709217.

54. Napolitano, F., Xu, X., and Gao, X. (2022). Impact of computational approaches in the fight against covid-19: an ai guided review of 17 000 studies. Briefings in Bioinformatics 23, bbab456. URL: https://academic.oup.com/bib/article/23/1/bbab456/6425232. doi: 10.1093/bib/bbab456.

55. Talevi, A., and Bellera, C.L. (2020). Challenges and opportunities with drug repurposing: finding strategies to find alternative uses of therapeutics. Expert Opinion on Drug Discovery 15, 397–401. URL: https://www.tandfonline.com/doi/full/10.1080/17460441.2020.1704729. doi: 10.1080/17460441.2020.1704729.

56. Shyam Sunder, S., Sunder, S.S., et al. (2023). Adverse effects of tyrosine kinase inhibitors in cancer therapy. Signal Transduction and Targeted Therapy 8, 262. URL: https://www.nature.com/articles/s41392-023-01469-6. doi: 10.1038/s41392-023-01469-6.

57. U.S. Food and Drug Administration (2023). Rinvoq (upadacitinib) prescribing information. URL: https://www.accessdata.fda.gov/drugsatfda_docs/label/2023/211675s015lbl.pdf boxed warning: serious infections, malignancy, major adverse cardiovascular events, thrombosis; Accessed 27 Jan 2026.

58. U.S. Food and Drug Administration (2022). Olumiant (baricitinib) prescribing information. URL: https://www.accessdata.fda.gov/drugsatfda_docs/label/2022/207924s006lbl.pdf boxed warning: serious infections, malignancy, major adverse cardiovascular events, thrombosis; Accessed 27 Jan 2026.

59. U.S. Food and Drug Administration (2025). Xeljanz / xeljanz xr / xeljanz oral solution (tofacitinib) prescribing information. URL: https://www.accessdata.fda.gov/drugsatfda_docs/label/2025/203214s038%2C208246s025%2C213082s010lbl.pdf boxed warning: serious infections, malignancy, major adverse cardiovascular events, thrombosis; Accessed 27 Jan 2026.

60. ClinicalTrials.gov (2024). Reverse-long covid-19 with baricitinib study (nct05858515). URL: https://clinicaltrials.gov/study/NCT05858515 phase 3 trial registration; Accessed 27 Jan 2026.

61. ClinicalTrials.gov (2025). Long covid (lc)-revitalize (nct06928272). URL: https://clinicaltrials.gov/study/NCT06928272 phase 3 trial registration; Accessed 27 Jan 2026.

62. ClinicalTrials.gov (2020). Early outpatient treatment for sars-cov-2 infection (covid-out) (nct04510194). URL: https://clinicaltrials.gov/study/NCT04510194 trial registration including metformin and ivermectin arms; Accessed 27 Jan 2026.

63. Geng, L.N., et al. (2024). Nirmatrelvir-ritonavir and symptoms in adults with postacute sequelae of sars-cov-2 infection: A randomized clinical trial. JAMA Internal Medicine. URL: https://jamanetwork.com/journals/jamainternalmedicine/fullarticle/2819901. Trial registration: NCT05576662.

64. ClinicalTrials.gov (2022). Study of nirmatrelvir/ritonavir for post-acute sequelae of covid-19 (stop-pasc) (nct05576662). URL: https://clinicaltrials.gov/study/NCT05576662 trial registration; Accessed 27 Jan 2026.

65. Pinero, S., Li, X., Liu, L., Li, J., Lee, S.H., Winter, M., Nguyen, T., Zhang, J., and Le, T.D. (2025). Integrative multi-omics framework for causal gene discovery in long covid. medRxiv pp. 2025.02.09.25321751. URL: https://www.medrxiv.org/content/10.1101/2025.02.09.25321751v1. doi: 10.1101/2025.02.09.25321751.

66. Pinero, S.L., Li, X., Liu, L., Li, J., Lee, S.H., Winter, M., Nguyen, T., Zhang, J., and Le, T.D. (2025). Taco: Tabpfn augmented causal outcomes for early detection of long covid. medRxiv pp. 2025.10.02.25337138. URL: https://www.medrxiv.org/content/10.1101/2025.10.02.25337138v1. doi: 10.1101/2025.10.02.25337138.

67. Wishart, D.S., Feunang, Y.D., Guo, A.C., Lo, E.J., Marcu, A., Grant, J.R., Sajed, T., Johnson, D., Li, C., Sayeeda, Z. et al. (2018). Drugbank 5.0: a major update to the drugbank database for 2018. Nucleic Acids Research 46, D1074–D1082. URL: 10.1093/nar/gkx1037. doi: 10.1093/nar/gkx1037.

68. Gaulton, A., Hersey, A., Nowotka, M., Bento, A.P., Chambers, J., Mendez, D., Mutowo, P., Atkinson, F., Bellis, L.J., Cibrián-Uhalte, E. et al. (2017). The chembl database in 2017. Nucleic Acids Research 45, D945–D954. URL: 10.1093/nar/gkw1074. doi: 10.1093/nar/gkw1074.

69. Wang, Y., Zhang, S., Li, F., Zhou, Y., Zhang, Y., Wang, Z., Zhang, R., Zhu, J., Ren, Y., Tan, Y. et al. (2020). Therapeutic target database 2020: enriched resource for facilitating research and early drug discovery. Nucleic Acids Research 48, D1031–D1041. URL: 10.1093/nar/gkz981. doi: 10.1093/nar/gkz981.

70. Ursu, O., Holmes, J., Knockel, J., Bologa, C.G., Yang, J.J., Mathias, S.L., Nelson, S.J., and Oprea, T.I. (2017). DrugCentral: online drug compendium. Nucleic Acids Research 45, D932–D939. doi: 10.1093/nar/gkw993.

71. Harding, S.D., Sharman, J.L., Faccenda, E., Southan, C., Pawson, A.J., Ireland, S., Gray, A.J.G., Bruce, L., Alexander, S.P.H., Anderton, S., Bryant, C., Davenport, A.P., Doerig, C., Fabbro, D., Levi-Schaffer, F., Spedding, M., Davies, J.A., and NC-IUPHAR (2018). The IUPHAR/BPS Guide to PHARMACOLOGY in 2018: updates and expansion to encompass the new Guide to IMMUNOPHARMACOLOGY. Nucleic Acids Research 46, D1091–D1106. doi: 10.1093/nar/gkx1121.

72. Falcon, S., and Gentleman, R. (2007). Using GOstats to test gene lists for GO term association. Bioinformatics 23, 257–258. doi: 10.1093/bioinformatics/btl567.

73. U.S. National Library of Medicine (2024). Clinicaltrials.gov. URL: https://clinicaltrials.gov/ accessed: 2025-07-25.

74. National Cancer Institute (). Common terminology criteria for adverse events (ctcae) v6.0 (meddra 28.0). URL: https://dctd.cancer.gov/research/ctep-trials/for-sites/adverse-events/ctcae-v6.pdf.

75. Kim, S., Thiessen, P.A., Bolton, E.E., Chen, J., Fu, G., Gindulyte, A., Han, L., He, J., He, S., Shoemaker, B.A., Wang, J., Yu, B., Zhang, J., and Bryant, S.H. (2016). Pubchem: a public information system for analyzing bioactivities of small molecules. Nucleic Acids Research 44, D1202–D1213. doi: 10.1093/nar/gkv951.

76. Kim, S., Thiessen, P.A., Bolton, E.E., and Bryant, S.H. (2018). Pubchem pug-rest: a restful web service for programmatic access to pubchem data. Nucleic Acids Research 46, W563–W570. doi: 10.1093/nar/gky294.

77. Gaulton, A. et al. (2012). Chembl: a large-scale bioactivity database for drug discovery. Nucleic Acids Research 40, D1100–D1107. doi: 10.1093/nar/gkr777.

78. Ochoa, D. et al. (2021). Open targets platform: supporting systematic drug–target identification and prioritisation. Nucleic Acids Research 49, D1302–D1310. doi: 10.1093/nar/gkaa1027.

79. U.S. National Library of Medicine (n.d.). Dailymed: Drug label information. National Library of Medicine (NLM).. Accessed programmatically via the DailyMed services API.

80. Lipinski, C.A., Lombardo, F., Dominy, B.W., and Feeney, P.J. (1997). Experimental and computational approaches to estimate solubility and permeability in drug discovery and development settings. Advanced Drug Delivery Reviews 23, 3–25. doi: 10.1016/S0169-409X(96)00423-1.

