## Supplementary material for "SPLIT: Safety Prioritization for Long COVID Drug Repurposing via a Causal Integrated Targeting Framework": SM

### **SPLIT: S**afety **P**rioritization for **L**ong COVID Drug Repurposing via a Causal **I**ntegrated **T**argeting Framework

Supplementary Materials

**Part II: Supplementary Results** 12

**9 Key Terms and Definitions** 12

**10 Causal Driver Gene Evidence** 13
10.1 Gene Counts by Method and Evidence Category . . . 13
10.2 Convergent Driver Genes . . . 13

**11 Drug–Gene Mapping Details** 13

**12 Drug Panel Threshold Justification** 14

**13 PlaNet Counterfactual Predictions** 16

**14 Adverse Event Signatures by Drug** 17

**15 Avoid List Structure and Concordance** 17
15.1 Avoid List Summary . . . 17
15.2 Concordance Metrics . . . 18

**16 Dimensional Risk Profiles** 18

**17 Secondary Screening: Drugs Failing Multiple Thresholds** 18

**18 Shared Deprioritized Drugs Across Cohorts** 19

19 Enrichment Analysis — Full Results 20

20 Supplementary Figures 20

### Part I: Supplementary Methods

#### 1 Causal Gene Discovery

To obtain a robust set of putative Long COVID driver genes, we combined three orthogonal causal inference approaches: Transcriptome-Wide Mendelian Randomization (TWMR), Control Theory (CT), and Differential Causal Effects (DCE). All methods were implemented using harmonized identifiers, consistent multiple-testing correction, and standardized quality-control procedures. The complete annotated gene list with evidence labels is provided in Supplementary Section S1.

##### 1.1 Transcriptome-Wide Mendelian Randomization (TWMR)

TWMR was used to identify genes whose genetically predicted expression levels exert a causal effect on Long COVID risk [6]. We integrated Genome-Wide Association Study (GWAS) summary statistics for Long COVID with quantitative trait locus of cis expression (cis-eQTL) data (GTEx v8) [16]. Genes were retained as drivers supported by TWMR when they showed: (i) consistent direction and magnitude among robust estimators, (ii) false discovery rate (FDR) $< 0.05$, and (iii) no evidence of substantial pleiotropic distortion based on heterogeneity and intercept tests. This step prioritizes targets with human genetic support, increasing the likelihood that modulating their expression affects Long COVID biology rather than reflecting confounding.

##### 1.2 Control Theory

To capture regulatory importance at the system level, we applied controllability analysis on a curated human protein–protein interaction network following Vinayagam *et al.* [22, 16]. We retained indispensable nodes whose removal increases the number of driver nodes required to control the network. Indispensable nodes have been reported to be enriched among essential genes, disease genes, and drug targets associated with severe adverse events [22, 15].

##### 1.3 Differential Causal Effects

To identify how regulatory pathways are causally altered in Long COVID, we implemented the DCE framework [7, 17]. RNA-seq data from individuals with Long COVID and recovered controls were analyzed within curated pathways (KEGG) to estimate directed causal effects between interacting genes separately in the Long COVID and control groups, while adjusting for confounders. We then computed differential causal effects across groups and ranked genes by their contribution to pathway perturbation. Genes with strong and consistent pathway reorganization signatures across multiple pathways were designated as DCE-supported drivers.

##### 1.4 Integration of Causal Evidence

We integrated the TWMR, CT, and DCE outputs to obtain a consolidated set of Long COVID driver genes. Genes supported by at least one method and not contradicted by sensitivity analyses were retained. This integration captures genetically anchored risk (TWMR), network-level regulatory leverage (CT), and condition-specific pathway reorganization (DCE).

#### 2 Clinical Trial Parsing and Knowledge Graph Integration

We implemented an automated workflow that (i) parses clinical trial records into structured trial-arm representations, (ii) resolves drugs and cohort descriptors to canonical biomedical identifiers, (iii) projects each arm into a typed-edge knowledge-graph representation, and (iv) supports counterfactual drug substitution while keeping the cohort and protocol constant.

##### 2.1 Trial Data Acquisition

Clinical trial records were retrieved from ClinicalTrials.gov using the public API (version 2). For each trial, we extracted the NCT identifier, arm group definitions, interventions, conditions, eligibility criteria, primary and secondary outcomes, phase, enrollment, and demographic constraints (sex and age ranges). Fields were normalized into a consistent internal schema for downstream parsing and graph construction. Raw clinical trial data for each cohort are provided in Supplementary Section S4 (NCT04809974) and Supplementary Section S5 (NCT04880161).

##### 2.2 Drug Entity Extraction and Standardization

Drug mentions in intervention descriptions were processed using MedEx [23] to extract medication entities and associated attributes when available. Extracted entities were mapped to canonical drug identifiers using a multi-stage strategy supporting exact matching, synonym lookup, and fuzzy matching with configurable similarity thresholds. Candidate replacement drugs were optionally validated against the PlaNet drug vocabulary prior to counterfactual generation; drugs not found were skipped.

##### 2.3 Eligibility Criteria Parsing and Population Characterization

Eligibility criteria were parsed using Criteria2Query [24] to extract structured inclusion/exclusion constraints. Extracted concepts were mapped to the Unified Medical Language System (UMLS) using concept search and Term Frequency-Inverse Document Frequency (TF–IDF) matching. Concepts were propagated through the UMLS hierarchy to enrich cohort representation with clinically relevant parent concepts.

##### 2.4 Outcome and Disease Normalization

Trial conditions were assigned to controlled disease identifiers, including Medical Subject Headings (MeSH) when available, to align trial context with the disease ontology represented in the knowledge graph. Primary outcomes were parsed and normalized to create structured outcome representations compatible with downstream feature encoding.

##### 2.5 Trial Arm Feature Encoding

For each arm, we constructed an arm text document by concatenating the intervention name/description, condition text, primary outcome text, arm label, trial attributes summary, and eligibility criteria text. In parallel, we derived structured trial attributes including phase encoding, logarithmic transformation of enrollment magnitude, sex eligibility, and age-range indicators. These attributes provide consistent low-dimensional covariates that complement text features in downstream embedding models. Parsed trial representations, including arm texts, eligibility criteria, and cohort descriptors, are provided in Supplementary Section S6 (NCT04809974) and Supplementary Section S7 (NCT04880161).

##### 2.6 Knowledge Graph Projection of Trial Arms

Each parsed trial arm was projected into the PlaNet KG by linking intervention, condition, population descriptors, and results to KG entities through typed relations. Arm indexing and labeling were resolved using multiple metadata fields and, when unavailable, by falling back to arm-group order. KG entity merges were handled using union-find over merge relations, and entity-to-KG-ID resolution used the PlaNet entity vocabulary.

##### 2.7 Counterfactual Drug Replacement and Leakage Controls

To evaluate candidate drugs within fixed cohort contexts, we generated counterfactual trial variants by substituting the experimental intervention while preserving all other protocol elements (condition, eligibility criteria, cohort descriptors, enrollment, demographics, and comparator structure). Replacement proceeded by identifying a non-placebo experimental intervention, using canonicalized names and identifiers when available. If a base trial lacked an explicit drug-annotated intervention, a minimal synthetic experimental anchor was created from the inferred non-placebo arm to allow replacement.

To reduce information leakage from real trial outcomes into counterfactual representations, counterfactual trials were cleaned by removing results-derived fields and replacing summaries with a neutral counterfactual statement. Arm labels and descriptions were updated to remove references to the original drug. These steps ensure that predictions are driven by cohort context and by the substituted intervention, rather than by accidental carryover from the original trial.

#### 3 PlaNet Model Architecture and Prediction Outputs

We applied PlaNet, a geometric deep learning framework trained on a large-scale clinical knowledge graph (KG), to predict cohort-anchored adverse-event profiles (AE), safety (S), and comparative efficacy (E) for each parsed trial arm [2]. In PlaNet, clinical trials are represented as trial-arm nodes connected via typed edges to protocol-defining entities, including administered drugs, investigated diseases/conditions, primary outcomes, and structured population descriptors derived from inclusion/exclusion criteria. This KG is integrated with biomedical background subnetworks (drug similarity/class hierarchies, disease hierarchies, drug–protein interactions, protein–protein interactions, and function ontologies), enabling message passing over higher-order relations among drugs, diseases, populations, and outcomes.

##### 3.1 PlaNet Encoder and Self-Supervised Pretraining

PlaNet learns low-dimensional embeddings for all KG entities using a relational graph convolutional network (R-GCN) encoder operating over typed edges. Entity representations are initialized from type-specific attributes and mapped into a shared embedding space via entity-type-specific linear projections; message passing then propagates information across multi-hop neighborhoods to produce final entity embeddings.

To leverage the large amount of unlabeled KG structure, PlaNet is first pretrained with a self-supervised link-prediction objective: random edges are masked or dropped from $k$-hop subgraphs and the model is trained to reconstruct missing edges using a bilinear scoring decoder (DistMult) over the learned embeddings. This pretraining encourages embeddings that capture both protocol-level trial structure and biomedical context, improving generalization to unseen drug–disease combinations.

##### 3.2 Trial-Arm Embedding Construction

For each trial arm, PlaNet constructs a protocol-level representation by combining (i) textual arm descriptors (arm title/description, intervention details, trial summary, eligibility criteria, and outcome text) embedded with a biomedical language model (BERT-family encoders), and (ii) structured trial covariates (phase, enrollment, age range, and sex eligibility). These arm attributes serve as input features for trial-arm nodes and are integrated with neighborhood information via the KG network, producing an embedding simultaneously conditioned on cohort context and tested intervention.

##### 3.3 Integrating New Trial Arms for Inference

For inference on newly parsed arms, including counterfactual substitutions, we instantiate trial-arm nodes with their arm text embeddings, structured covariates, and KG edges (drug, disease, outcome, and population links). Predictions are then obtained by applying the pretrained PlaNet encoder and corresponding task heads to the resulting arm embeddings, without re-training.

##### 3.4 Adverse Event and Safety Prediction

PlaNet's safety and adverse-event tasks are defined relative to a placebo (no-treatment) baseline for the same disease and population context. Placebo arms are aggregated to estimate a cohort-anchored prior probability of adverse events for each disease and associated population, representing expected events in the absence of active intervention. Given an intervention arm, PlaNet predicts whether (and how strongly) an adverse event is enriched compared to this placebo prior, implemented via contingency-table comparisons against the estimated placebo distribution and an odds-ratio enrichment threshold [2].

We query two PlaNet safety-related prediction heads: (i) a binary serious adverse event (SAE) head that outputs a probability of SAE occurrence or enrichment for a given arm; and

(ii) a multi-task AE-category head that outputs calibrated probabilities over a standardized adverse-event vocabulary defined at the Medical Dictionary for Regulatory Activities (MedDRA) Preferred Term (PT) level [11]. For each arm $i$ and adverse-event category $k$, PlaNet produces:

$$p_{i,k} = \Pr(\text{AE category } k \text{ is enriched in arm } i \text{ relative to placebo}) \in [0, 1].$$

To align with our interpretation where higher values indicate safer profiles, we define the global safety score as the complement of SAE risk:

$$S_{\text{raw},i} = 1 - p_i^{\text{SAE}},$$

where $p_i^{\text{SAE}}$ is the SAE-head probability for arm $i$. A larger $S_{\text{raw}}$ indicates safer predicted profiles, while a larger $p_{i,k}$ indicates higher predicted risk of AE enrichment for category $k$.

##### 3.5 Comparative Efficacy Prediction

PlaNet's efficacy task is trained on survival-type endpoints extracted from completed clinical trials. For trials containing two or more arms with comparable outcome definitions, PlaNet is fine-tuned to predict which arm is expected to achieve a more favorable survival outcome. When a placebo or comparator arm could be reliably identified, we computed cohort-specific comparative efficacy as:

$$E_{\text{raw}} = \Pr(\text{active arm is superior to comparator}), \qquad E_{\text{raw}} \in [0, 1].$$

Values above 0.5 indicate higher predicted probability of benefit under the active intervention, conditional on cohort and protocol context encoded in the trial-arm representation. When placebo identification was ambiguous or a valid pairing could not be established, efficacy was not calculated, and subsequent evaluation relied only on safety and adverse-event components.

##### 3.6 Cohort-Anchored Difference Metrics

For each metric $X \in \{S_{\text{raw}}, AE_{\text{score}}, E_{\text{raw}}\}$, the counterfactual delta is defined as:

$$\Delta X = X_{\text{counterfactual}} - X_{\text{reference}}.$$

Negative $\Delta S$ or $\Delta E$ indicates a worse profile under substitution; positive $\Delta AE$ indicates increased toxicity burden.

### 4 AE Burden Scoring and Weighting

#### 4.1 Weighted Adverse-Event Burden Score

To summarize toxicity while prioritizing clinically severe and Long COVID-relevant events, we computed a weighted probability for each event $k$ in arm $i$:

$$\hat{p}_{i,k} = p_{i,k} \cdot s_k \cdot \ell_k,$$

where $s_k$ is a CTCAE-aligned severity weight and $\ell_k$ is a Long COVID relevance weight. The weighted AE burden score is then:

$$AE_{\text{score},i} = \sum_k \hat{p}_{i,k}.$$

Larger values indicate worse predicted AE burden.

#### 4.2 Severity Weights

Severity weights ($s_k$) were assigned based on CTCAE-aligned severity categories [11]:

| **Severity Category** | **Weight ($s_k$)** |
|---|---|
| Mild | 0.25 |
| Moderate | 0.50 |
| Severe | 0.75 |
| Life-threatening | 1.00 |
| Death | 1.20 |

Table 1: CTCAE-aligned severity weights for adverse event scoring.

#### 4.3 Long COVID Relevance Weights

Relevance weights ($\ell_k$) were assigned based on clinical relevance to Long COVID symptom domains:

| **Relevance Category** | **Weight ($\ell_k$)** |
|---|---|
| Core Long COVID symptoms | 1.50 |
| Important comorbidity | 1.25 |
| General | 1.00 |

Table 2: Long COVID relevance weights for adverse event scoring.

Core Long COVID symptoms include fatigue, cognitive dysfunction ("brain fog"), dyspnea, chest pain, palpitations, postural symptoms, and persistent anosmia/dysgeusia. Important comorbidities include cardiovascular events, thromboembolic events, and metabolic disturbances frequently observed in Long COVID populations.

When a weight could not be confidently assigned, default values of $s_k = 0.50$ and $\ell_k = 1.00$ were applied.

### 5 Composite Ranking Equations and Normalization

#### 5.1 Min–Max Normalization

For cohort-internal ranking, raw scores were normalized using min–max scaling within each cohort:

$$\tilde{X} = \frac{X_{\text{raw}} - X_{\text{min}}}{X_{\text{max}} - X_{\text{min}}},$$

where $X \in \{S, E\}$ and $X_{\text{min}}$, $X_{\text{max}}$ are the minimum and maximum values observed within the cohort.

#### 5.2 Inverse AE Burden Normalization

To convert AE burden (where larger = worse) into a scale compatible with safety and efficacy (where larger = better), we computed the inverse AE burden:

$$AE^{-1} = \frac{1}{AE_{\text{score}} + \epsilon}, \qquad \epsilon = 10^{-9},$$

followed by cohort-level min–max normalization:

$$\widehat{AE^{-1}} = \frac{AE^{-1} - (AE^{-1})_{\text{min}}}{(AE^{-1})_{\text{max}} - (AE^{-1})_{\text{min}}}.$$

#### 5.3 Composite Score Computation

The three components, normalized safety ($\tilde{S}$), normalized efficacy ($\tilde{E}$), and normalized inverse AE burden ($\widehat{AE^{-1}}$), were combined into a single composite ranking score:

$$C = w_S \cdot \tilde{S} + w_E \cdot \tilde{E} + w_{AE} \cdot \widehat{AE^{-1}},$$

with default weights $w_S = 0.4$, $w_E = 0.4$, and $w_{AE} = 0.2$, constrained such that $w_S + w_E + w_{AE} = 1$. These weights reflect the safety-first principle of SPLIT while treating efficacy as a co-dominant signal. The weight assigned to S is equal to that of E, with AE burden receiving a lower weight because it is partially captured by the safety score.

When comparative efficacy ($E$) was unavailable for a given cohort, weights were renormalized over the remaining components:

$$C = \frac{w_S}{w_S + w_{AE}} \cdot \tilde{S} + \frac{w_{AE}}{w_S + w_{AE}} \cdot \widehat{AE^{-1}} = 0.67 \cdot \tilde{S} + 0.33 \cdot \widehat{AE^{-1}}.$$

### 6 Avoid Lists: Definitions, Thresholds, and Stability

#### 6.1 Primary Composite Avoid List

Drugs were ranked by composite score $C$, with lowest-scoring candidates forming the avoid list. We report three stringency levels: q05 (worst 5%), q10 (worst 10%, default), and q20 (worst 20%).

##### 6.2 Secondary Threshold-Based Screening

As an independent safety check, we flagged drugs with extreme individual metrics (q20 thresholds): AE-extreme (top 20% of $AE_{\text{score}}$), safety-extreme (bottom 20% of $S_{\text{raw}}$), and efficacy-extreme (bottom 20% of $E_{\text{raw}}$). We tracked the number of triggered flags per drug ($n_{\text{extremes}} \in \{1, 2, 3\}$).

##### 6.3 Counterfactual-Delta Avoid List

We identified drugs performing worse than the original intervention under substitution. Candidates were flagged as delta-worse when $\Delta S < 0$, $\Delta E < 0$, or $\Delta AE > 0$. The delta avoid list includes candidates in the worst decile for any metric, with multi-flag tracking ($n_{\text{delta}} \in \{1, 2, 3\}$).

##### 6.4 Cross-List Concordance and Sensitivity Analysis

Candidates appearing across multiple lists (composite, threshold, and delta) represent higher-confidence deprioritization targets. We assessed sensitivity by repeating analyses across q05, q10, and q20 thresholds. Candidates stable across thresholds form a conservative "avoid" core; threshold-dependent candidates define a broader caution set.

#### 7 Rule-Based Filtering Categories

##### 7.1 Hard-Block Rules

Candidates were automatically excluded if they matched any of the following criteria:

- Oncology-specific agents (chemotherapeutics, targeted cancer therapies) without established non-oncology indications
- Acute-only interventions (e.g., thrombolytics, acute sedatives) unsuitable for chronic/outpatient Long COVID management
- Withdrawn or revoked drugs
- Agents with unacceptable safety profiles for the target population (e.g., known teratogens when reproductive-age populations are included)

##### 7.2 Soft-Block Rules

Candidates were flagged for manual review (but not automatically excluded) if they:

- Require intensive monitoring (e.g., narrow therapeutic index, mandatory lab monitoring)
- Have significant drug–drug interaction potential relevant to common Long COVID comedications
- Are associated with known risks in Long COVID-relevant organ systems (cardiac, pulmonary, neurological) but may still offer net benefit
- Have limited safety data in post-viral or chronic fatigue populations

##### 7.3 Keep-Hint Categories

Candidates were tagged for preserved visibility if they aligned with Long COVID-relevant therapeutic themes:

- Cardiopulmonary/vascular: Agents addressing cardiovascular symptoms, microvascular dysfunction, or endothelial health
- Respiratory anti-inflammatory: Inhaled or systemic anti-inflammatory agents for persistent respiratory symptoms
- Neurocognitive: Agents targeting cognitive dysfunction, neuroinflammation, or autonomic dysregulation
- Metabolic/fibrotic: Agents addressing metabolic dysregulation or fibrotic sequelae
- Autonomic/POTS-related: Agents for postural orthostatic tachycardia syndrome and related dysautonomia
- Low-risk supportive: Well-tolerated supportive agents (e.g., supplements, low-risk symptomatic treatments)

### 8 Pharmacology Enrichment Protocol

##### 8.1 Data Sources

Enrichment data were collected programmatically from:

- PubChem [9, 8]: Cross-references, physicochemical properties, compound identifiers
- ChEMBL [4]: Potency/binding data ($IC_{50}$, $EC_{50}$, $K_i$, $K_d$), mechanism of action, target annotations
- Open Targets [13]: Disease associations, target tractability, clinical precedence
- DailyMed [21]: FDA label information, boxed warnings, contraindications

##### 8.2 Physicochemical Properties Collected

- Molecular weight (MW)
- Lipophilicity (XLogP, ALogP)
- Topological polar surface area (TPSA)
- Hydrogen bond donors (HBD)
- Hydrogen bond acceptors (HBA)
- Rotatable bond count (flexibility)
- Heavy atom count

##### 8.3 Lipinski Rule-of-Five Assessment

Developability was assessed using the Lipinski Rule-of-Five [10]. Violations were counted for:

- MW $>$ 500 Da
- XLogP $> 5$
- HBD $> 5$
- HBA $> 10$

Compounds with $\leq 1$ violation were considered to have favorable oral bioavailability profiles.

### Part II: Supplementary Results

#### 9 Key Terms and Definitions

For clarity, we define key terms used throughout the main text and supplementary materials:

- **Prediction**: PlaNet outputs for S, E, and AE probabilities in a given cohort context.
- **Counterfactual substitution**: Replacing the trial intervention while maintaining the cohort, protocol, and comparator fixed.
- **Evaluation**: The process of testing SPLIT as a framework, using counterfactual substitution to assess whether predicted scores distinguish drugs with different risk profiles.
- **Screening**: Applying predictions and scoring across a drug panel to generate ranks.
- **AE burden**: A weighted score combining individual predicted AE probabilities with severity and Long COVID relevance weights, summarizing the overall predicted toxicity load of a candidate.
- **Composite score**: A single ranking metric that combines normalized S, E, and inverse AE burden to summarize the overall risk-benefit profile of each candidate.
- **Deprioritization**: Labeling high-risk or low-benefit candidates as "avoid" based on ranks, thresholds, or deltas relative to the original trial drug.
- **Template cohort**: The primary clinical trial context (NCT04809974, cognitive phenotype) in which counterfactual substitutions are performed.
- **Sensitivity cohort**: A second clinical trial context (NCT04880161, respiratory phenotype) used to test whether deprioritization patterns generalize across phenotypes.

### 10 Causal Driver Gene Evidence

#### 10.1 Gene Counts by Method and Evidence Category

Table 3 summarizes the distribution of causal driver genes across the three inference methods. CT contributed the largest set (1,641 genes), reflecting the scale of the protein–protein interaction network, while TWMR (50 genes) and DCE (45 genes) provided complementary, more focused evidence.

Table 3: **Summary of causal driver gene evidence across methods:** Mendelian Randomization (MR), Control Theory (CT), and Differential Causal Effects (DCE). Full gene list in Supplementary Section S1.

| Evidence category | Genes (n) |
|---|---|
| Total causal driver genes | 1,725 |
| CT supported (any) | 1,641 |
| MR supported (any) | 50 |
| DCE supported (any) | 45 |
| ***Single method support*** | |
| CT only | 1,631 |
| MR only | 45 |
| DCE only | 38 |
| ***Support from multiple methods*** | |
| CT + DCE | 6 |
| CT + MR | 4 |
| DCE + MR | 1 |
| Convergent total | 11 |

#### 10.2 Convergent Driver Genes

Table 4 lists all 11 genes supported by two or more causal inference methods.

Table 4: **Long COVID causal driver genes supported by ≥2 causal inference methods.** Complete annotations in Supplementary Section S1.

| Gene | Supporting methods |
|---|---|
| *ERCC3* | CT; MR |
| *HIF1A* | CT; DCE |
| *IMMT* | CT; MR |
| *LEF1* | CT; DCE |
| *MED19* | CT; MR |
| *MYC* | CT; DCE |
| *NDUFA6* | CT; MR |
| *PNPO* | DCE; MR |
| *POMC* | CT; DCE |
| *STAT3* | CT; DCE |
| *TP53* | CT; DCE |

### 11 Drug–Gene Mapping Details

Table 5 presents the 10 drugs with the highest causal gene coverage. The dominance of oncology kinase inhibitors in this list motivated the explicit deprioritization step in the framework. Complete mapping tables are provided in Supplementary Section S2.

Table 5: **Top 10 drugs by number of mapped Long COVID causal gene targets.** Full mapping table in Supplementary Section S2.

| Drug | CG (n) | CG Methods | Drug–target sources |
|---|---|---|---|
| Imatinib | 210 | CT; DCE | ChEMBL; DrugBank; DrugCentral; GtoPdb |
| Sirolimus | 206 | CT; DCE | ChEMBL; DrugCentral; GtoPdb |
| Palbociclib | 201 | CT | ChEMBL; DrugBank; DrugCentral; GtoPdb; TTD |
| Sunitinib | 194 | CT | ChEMBL; DrugBank; DrugCentral; GtoPdb |
| Sorafenib | 191 | CT; DCE | ChEMBL; DrugBank; DrugCentral; GtoPdb; TTD |
| Nilotinib | 190 | CT | ChEMBL; DrugBank; DrugCentral; GtoPdb |
| Bosutinib | 189 | CT; DCE | ChEMBL; DrugBank; DrugCentral; GtoPdb; TTD |
| Gefitinib | 189 | CT | ChEMBL; DrugBank; DrugCentral; GtoPdb; TTD |
| Erlotinib | 186 | CT | ChEMBL; DrugBank; DrugCentral; GtoPdb; TTD |
| Vandetanib | 185 | CT | ChEMBL; DrugBank; DrugCentral; GtoPdb; TTD |

**Abbreviations:** n, Number; CG, Causal Gene Targets; CT, Control Theory; DCE, Differential Causal Effects.

### 12 Drug Panel Threshold Justification

To address potential circularity in threshold selection, we independently derived $k \geq 11$ via a hypergeometric enrichment test that asks, for each of the 19,172 candidate drugs, whether its overlap with the 1,725 Long COVID causal genes is greater than expected by chance against a background of 20,000 human genes [3], with Benjamini-Hochberg correction applied across all drugs.

Four findings collectively justify $k = 11$ as the principal threshold (Figure 1). First, the distribution of gene targets across 19,172 drugs is highly right-skewed: the 75th percentile is only 2 genes, which means $k = 11$ already exceeds the 90th percentile of the complete distribution (Figure 1A). Second, at $k = 11$, the hypergeometric test confirms that 100% of retained drugs have a statistically non-random overlap with the causal gene network (FDR $< 0.05$; Figure 1B). This proportion drops to 98.1% at $k = 10$ and to 75.4% at $k = 5$, demonstrating that $k = 11$ marks a sharp transition point at which enrichment significance becomes complete and stable. Third, sensitivity analysis shows that the panel size decreases smoothly from 1,776 drugs at $k = 11$ to 1,378 at $k = 20$ and 1,065 at $k = 30$ (Figure 1C), without discontinuous collapse, indicating that deprioritization results are robust to modest variation in threshold. Fourth, the integration of all three causal inference methods is a prerequisite for a tractable panel at this threshold: at $k = 11$, using MR genes alone or DCE genes alone produces panels of zero drugs, while CT alone produces 991 drugs, substantially below the 1,776 obtained from the union of CT, MR, and DCE (Figure 1D). This result directly supports the methodological decision to integrate three complementary causal inference approaches rather than relying on any single method.

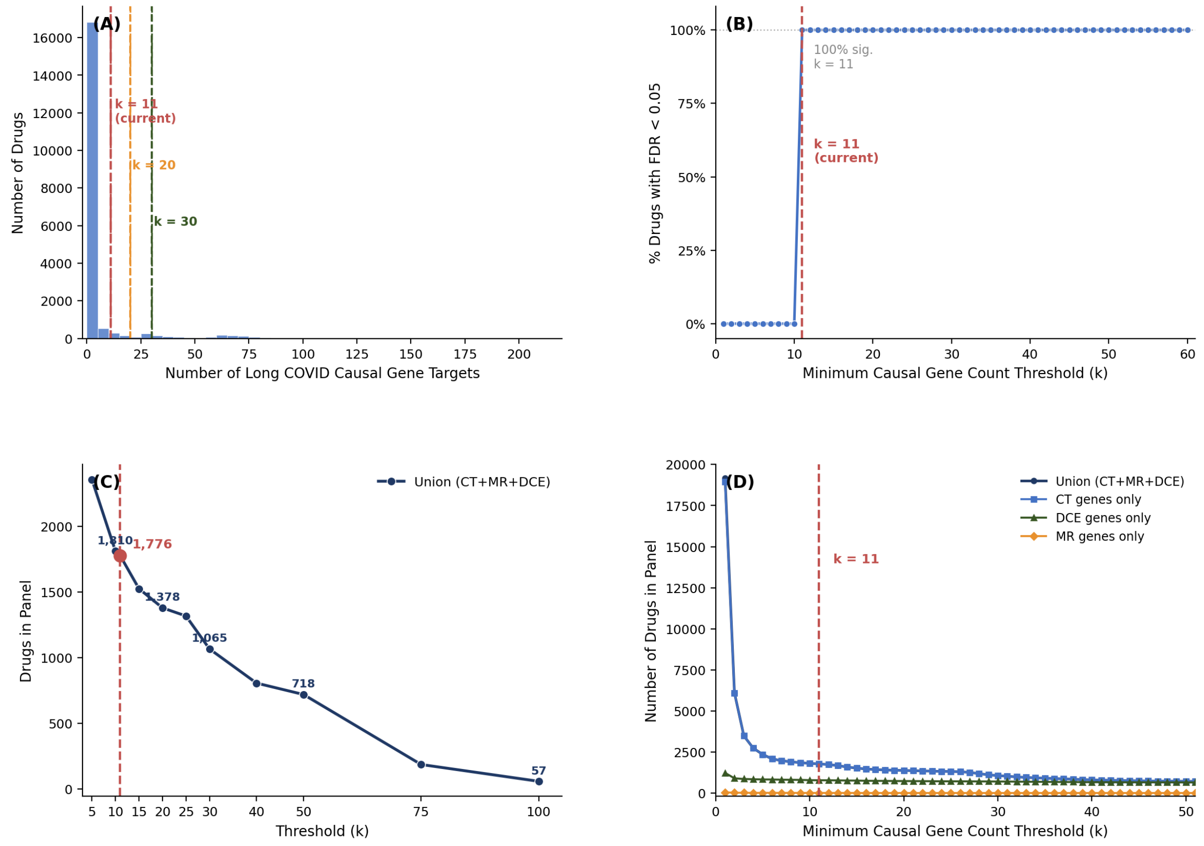

Figure 1: **Threshold justification for the screening panel.** (A) Distribution of causal gene targets per drug across all 19,172 candidates. The distribution is highly right-skewed; the 75th percentile is 2 genes and $k = 11$ exceeds the 90th percentile. Vertical dashed lines indicate thresholds $k = 11$ (red), $k = 20$ (orange), and $k = 30$ (green). (B) Percentage of drugs with statistically significant causal gene enrichment (FDR $< 0.05$, hypergeometric test with BH correction against a 20,000-gene background) as a function of minimum gene count threshold. At $k = 11$, enrichment significance reaches 100% and remains stable at all higher thresholds. (C) Panel size as a function of threshold. The smooth decrease from 1,776 ($k = 11$) to 57 ($k = 100$) confirms that the threshold selection does not produce a discontinuous cut. (D) Panel size stratified by gene source (CT only, MR only, DCE only, and union CT+MR+DCE). At $k = 11$, MR-only and DCE-only panels are empty, confirming that the union of all three methods is necessary to obtain a tractable screening panel. Red dashed line in all panels: $k = 11$ (current threshold). Background $N = 20{,}000$ genes; $K = 1{,}725$ Long COVID driver genes. FDR: Benjamini-Hochberg.

We also tested whether the choice of threshold affects the safety profile of the retained panel. Across thresholds from $k = 11$ to $k = 50$, the safety score distribution is stable in both cohorts (Figure 2). In the cognitive cohort, the proportion of drugs with $S \geq 0.4$ ranges from 9% to 11% at all tested thresholds; in the respiratory cohort, from 73% to 77% (Figure 2A). The full safety score distributions, shown as box plots, confirm that neither the interquartile range nor the median shifts meaningfully across thresholds in either cohort (Figure 2B). The median S in the cognitive cohort decreases only marginally from 0.29 at $k = 11$ to 0.25 at $k = 50$ (Figure 2C), while panel size halves over the same range.

This stability is a direct structural consequence of the safety-coverage trade-off described in the main text ($r = -0.99$). Higher thresholds preferentially retain drugs with broader causal gene coverage, which are precisely the oncology kinase inhibitors with the highest AE burden and the lowest predicted S. The enrichment of high-coverage drugs therefore exactly offsets any improvement in panel selectivity, leaving the safety distribution unchanged. The flat safety sensitivity curves are thus a quantitative signature of that trade-off operating at the panel level. Taken together, these analyses confirm that $k = 11$ is statistically principled, that it is not sensitive to modest variation, and that raising it would reduce panel coverage without improving the safety profile.

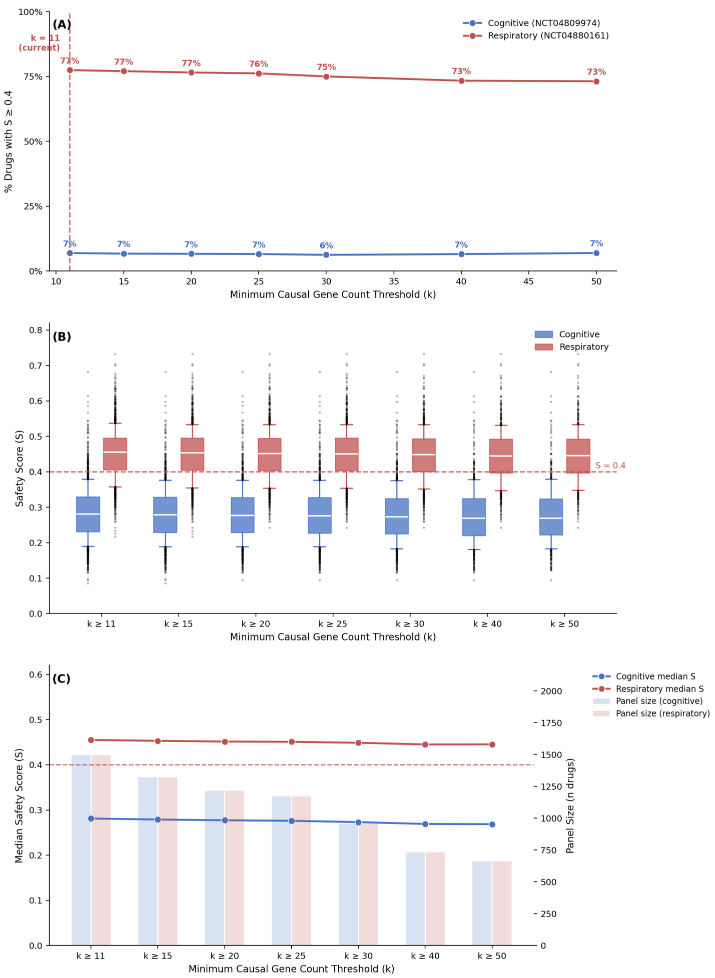

Figure 2: **Safety score distribution is stable across gene count thresholds.** (A) Percentage of drugs with $S \geq 0.4$ (SPLIT safety threshold) at each minimum gene count threshold from $k = 11$ to $k = 50$, for both cohorts. Cognitive cohort (blue): 9–11%; respiratory cohort (red): 73–77%. (B) Box plots of the full safety score distribution at each threshold. Box: IQR; whiskers: 10th–90th percentiles; dots: outliers. Red dashed line: $S = 0.4$ safety threshold. (C) Median S (lines, left axis) and panel size (bars, right axis) as a function of threshold. As panel size decreases from approximately 1,600 at $k = 11$ to approximately 500 at $k = 50$, median S changes by at most 0.04 (cognitive) and 0.02 (respiratory). The flat safety sensitivity curves are a structural consequence of the safety-coverage trade-off ($r = -0.99$): higher thresholds enrich for high-coverage kinase inhibitors, which are the least safe drugs, canceling any enrichment effect on the safety distribution. Red dashed line: $k = 11$ (current threshold).

### 13 PlaNet Counterfactual Predictions

Table 6 provides the full PlaNet predictions for the template cohort (NCT04809974), including the original trial drug (Niagen) and three counterfactual substitutions. These data underlie the multi-panel Figure 2 in the main text. Complete prediction outputs are provided in Supplementary Section S8.

Table 6: **PlaNet predictions for the NCT04809974 template cohort, original trial drug (Niagen), and counterfactual substitutions.** S and AE outputs are defined relative to a placebo baseline in the same population context, whereas the Δ values quantify differences between candidates within the cohort (candidate minus Niagen). S scores range from 0 to 1 (higher = safer); E represents $E = \Pr(\text{Drug} > \text{Placebo})$. Top AE is the highest probability event. Full AE profiles in Supplementary Section S3.

| Drug | CG | S | $\Delta S$ | E | $\Delta E$ | Top AE (P) |
|---|---|---|---|---|---|---|
| Niagen (Original) | — | 0.549 | — | 0.519 | — | Hypertensive diseases (0.197) |
| Leronlimab | 1 | 0.441 | −0.108 | 0.521 | +0.002 | Hypertensive diseases (0.175) |
| Vortioxetine | 11 | 0.332 | −0.217 | 0.521 | +0.002 | Nausea (0.234) |
| Ritonavir | 70 | 0.215 | −0.334 | 0.527 | +0.009 | Diarrhea (0.192) |

**Abbreviations:** CG, number of causal genes targeted; S, safety score; E, efficacy probability; AE, adverse event; P, probability.

### 14 Adverse Event Signatures by Drug

Table 7 presents the top 10 predicted AE for each counterfactual scenario in the NCT04809974 cohort, enabling distinction between drug-related toxicity and cohort background signals. Complete probability tables are provided in Supplementary Section S3, with mapped AE tables in Supplementary Section S10.

Table 7: **Top 10 predicted adverse events per scenario in the NCT04809974 cohort.** Probabilities represent PlaNet predictions. Full AE tables in Supplementary Section S3.

| Rank | Niagen (Original) | P | Rank | Ritonavir | P |
|---|---|---|---|---|---|
| 1 | Hypertensive diseases | 0.197 | 1 | Diarrhoea | 0.192 |
| 2 | Diarrhoea | 0.174 | 2 | Hypertensive diseases | 0.136 |
| 3 | Confusion NOS | 0.173 | 3 | Acrodermatitis | 0.125 |
| 4 | Bronchopneumonia | 0.157 | 4 | Hypokalaemia | 0.100 |
| 5 | Hypokalaemia | 0.139 | 5 | Asthenia | 0.099 |
| 6 | Hyperglycaemia | 0.122 | 6 | Dyspnoea | 0.095 |
| 7 | Nausea with vomiting | 0.118 | 7 | Bronchopneumonia | 0.084 |
| 8 | Asthenia | 0.115 | 8 | Confusion NOS | 0.083 |
| 9 | Nausea | 0.113 | 9 | Dehydration | 0.082 |
| 10 | Dyspnoea | 0.111 | 10 | Headache | 0.075 |
| **Rank** | **Vortioxetine** | **P** | **Rank** | **Leronlimab** | **P** |
| 1 | Nausea | 0.234 | 1 | Hypertensive diseases | 0.175 |
| 2 | Confusion NOS | 0.182 | 2 | Asthenia | 0.157 |
| 3 | Diarrhoea | 0.177 | 3 | Diarrhoea | 0.144 |
| 4 | Nausea with vomiting | 0.168 | 4 | Hyperglycaemia | 0.137 |
| 5 | Insomnia | 0.162 | 5 | Dyspnoea | 0.126 |
| 6 | Constipation | 0.155 | 6 | Headache | 0.124 |
| 7 | Hypertensive diseases | 0.148 | 7 | Bronchopneumonia | 0.118 |
| 8 | Anorexia | 0.142 | 8 | Acrodermatitis | 0.118 |
| 9 | Asthenia | 0.141 | 9 | Hypokalaemia | 0.111 |
| 10 | Weight below normal | 0.089 | 10 | Confusion NOS | 0.111 |

### 15 Avoid List Structure and Concordance

#### 15.1 Avoid List Summary

Table 8 summarizes the structure and output of each deprioritization category for both cohorts.

Table 8: **Summary of deprioritization avoid lists for both cohorts.** Quantile thresholds (q05, q10, q20) indicate the percentage of worst ranked drugs captured. NCT04809974 deprioritization is driven by S/AE; NCT04880161 deprioritization is driven by E. Full avoid lists in Supplementary Section S14 (NCT04809974) and Supplementary Section S15 (NCT04880161).

| List type | Threshold | C1 | C2 | Description |
|---|---|---|---|---|
| ***Primary composite (worst overall risk–benefit)*** | | | | |
| Primary q05 | Worst 5% | 82 | 82 | Core deprioritized set |
| Primary q10 | Worst 10% | 163 | 163 | Default threshold |
| Primary q20 | Worst 20% | 325 | 325 | Extended set |
| ***Secondary (extreme individual metrics)*** | | | | |
| Threshold q20 | Any extreme | 855 | 803 | Drugs with ≥1 extreme flag |
| AE extreme | Top 20% AE | 326 | 326 | High predicted toxicity burden |
| Safety extreme | Bottom 20% S | 325 | 325 | Low predicted S score |
| Efficacy extreme | Bottom 20% E | 325 | 325 | Low predicted E |
| Multiple extremes | ≥2 flags | 121 | 168 | Failed multiple thresholds |
| ***Counterfactual delta (worse than original trial drug)*** | | | | |
| Delta q10 | Any worse | 457 | 412 | Drugs worse than original |
| ΔAE worse | – | 163 | 163 | Higher AE burden |
| ΔS worse | – | 163 | 163 | Lower S |
| ΔE worse | – | 164 | 163 | Lower E |
| ***Concordance across lists (q10 primary)*** | | | | |
| Primary ∩ Secondary | – | 158 (96.9%) | 162 (99.4%) | Overlap with threshold list |
| Primary ∩ Delta | – | 113 (69.3%) | 131 (80.4%) | Overlap with delta list |
| All three lists | – | 113 | 131 | Highest confidence targets |

**Abbreviations:** C1, Cohort 1 (NCT04809974); C2, Cohort 2 (NCT04880161). Original trial drugs are Niagen (NCT04809974) and Ampion (NCT04880161). Panel sizes: NCT04809974 = 1,625 drugs; NCT04880161 = 1,630 drugs.

##### 15.2 Concordance Metrics

Table 9 provides detailed concordance metrics across avoid list categories.

Table 9: **Concordance across avoid lists.** The respiratory cohort shows higher concordance across all metrics, reflecting more consistent deprioritization signals when E provides strong discrimination.

| Concordance metric | NCT04809974 | NCT04880161 | Difference |
|---|---|---|---|
| Primary list size | 163 (10.0%) | 163 (10.0%) | — |
| Secondary list size | 855 (52.6%) | 803 (49.4%) | −3.2% |
| Delta list size | 457 (28.1%) | 412 (25.4%) | −2.7% |
| Primary ∩ Secondary | 158 (96.9%) | 162 (99.4%) | +2.5% |
| Primary ∩ Delta | 113 (69.3%) | 131 (80.4%) | +11.1% |
| All three lists | 113 | 131 | +18 drugs |

#### 16 Dimensional Risk Profiles

Table 10 presents drugs with the most extreme unfavorable values on each individual risk dimension, revealing how each metric discriminates differently across the two cohorts.

#### 17 Secondary Screening: Drugs Failing Multiple Thresholds

Table 11 presents the top 10 drugs failing multiple thresholds in both cohorts. Oncology kinase inhibitors dominated this subset (70% of flagged drugs), with the respiratory cohort showing 2–3× higher AE values and lower E.

Table 10: **Bottom 5 drugs by individual risk dimension for both cohorts.** E shows minimal discrimination in the cognitive cohort (spread = 0.002) but strong discrimination in the respiratory cohort (spread = 0.015 among worst 10). AE burden is approximately 2× higher in the respiratory cohort. Full rankings in Supplementary Section S12 and Supplementary Section S13.

| NCT04809974 (Cognitive) | | | | NCT04880161 (Respiratory) | | | |
|---|---|---|---|---|---|---|---|
| ***Worst by AE Burden*** | | | | | | | |
| **R** | **Drug** | **AE** | **Class** | **R** | **Drug** | **AE** | **Class** |
| 1 | Lenvatinib | 14.97 | Multi-kinase | 1 | Cobimetinib | 30.83 | MEK inh. |
| 2 | Carfilzomib | 14.85 | Proteasome | 2 | Carfilzomib | 30.11 | Proteasome |
| 3 | Cobimetinib | 14.72 | MEK inh. | 3 | Lumacaftor | 29.88 | CFTR corr. |
| 4 | Regorafenib | 14.24 | Multi-kinase | 4 | Selumetinib | 28.92 | MEK inh. |
| 5 | Axitinib | 13.72 | VEGFR inh. | 5 | Lenvatinib | 28.85 | Multi-kinase |
| *Range: 12.8–15.0* | | | | *Range: 25.2–30.8 (2× higher)* | | | |
| ***Worst by Safety*** | | | | | | | |
| **R** | **Drug** | **S** | **Class** | **R** | **Drug** | **S** | **Class** |
| 1 | Benzoyl peroxide | 0.086 | Topical | 1 | Benzoyl peroxide | 0.217 | Topical |
| 2 | Terconazole | 0.094 | Topical antifung. | 2 | Glucosamine | 0.226 | Nutraceutical |
| 3 | Cetylpyridinium | 0.097 | Topical antisep. | 3 | Cetylpyridinium | 0.233 | Topical antisep. |
| 4 | Donepezil | 0.116 | AChE inh. | 4 | Salsalate | 0.243 | NSAID |
| 5 | Scopolamine | 0.116 | Anticholinergic | 5 | Folic acid | 0.259 | Vitamin |
| *Range: 0.09–0.12 (very low)* | | | | *Range: 0.22–0.27 (higher floor)* | | | |
| ***Worst by Efficacy*** | | | | | | | |
| **R** | **Drug** | **E** | **Class** | **R** | **Drug** | **E** | **Class** |
| 1 | Pivampicillin | 0.514 | Penicillin | 1 | Olaparib | 0.009 | PARP inh. |
| 2 | Upadacitinib | 0.515 | JAK inh. | 2 | Temozolomide | 0.014 | Alkylating |
| 3 | Chlorphentermine | 0.515 | Anorectic | 3 | Enzalutamide | 0.015 | Antiandrogen |
| 4 | Bithionol | 0.515 | Antiparasitic | 4 | Alectinib | 0.017 | ALK inh. |
| 5 | Estradiol cyp. | 0.515 | Estrogen | 5 | Trametinib | 0.019 | MEK inh. |
| *Range: 0.514–0.516 (no discrim.)* | | | | *Range: 0.009–0.024 (strong discrim.)* | | | |

**Abbreviations:** R, rank; inh., inhibitor; corr., corrector; antifung., antifungal; antisep., antiseptic; cyp., cypionate; discrim., discrimination.

Table 11: **Top 10 drugs failing multiple thresholds in both cohorts.** All listed drugs were flagged on both AE burden and E. Oncology kinase inhibitors dominate both lists, but failure modes differ quantitatively: the respiratory cohort shows 2–3× higher AE values and lower E. Full screening in Supplementary Section S14 and Supplementary Section S15.

| NCT04809974 (Cognitive) | | | | | NCT04880161 (Respiratory) | | | | |
|---|---|---|---|---|---|---|---|---|---|
| **R** | **Drug** | **AE** | **S** | **E** | **R** | **Drug** | **AE** | **S** | **E** |
| 1 | Cobimetinib | 5.5 | .51 | .519 | 1 | Carfilzomib | 13.0 | .37 | .045 |
| 2 | Regorafenib | 5.2 | .55 | .517 | 2 | Selumetinib | 12.0 | .35 | .029 |
| 3 | Infigratinib | 4.6 | .38 | .519 | 3 | Cobimetinib | 11.8 | .41 | .039 |
| 4 | Ruxolitinib | 4.6 | .53 | .519 | 4 | Pomalidomide | 11.6 | .37 | .053 |
| 5 | Tazemetostat | 4.3 | .36 | .519 | 5 | Lenvatinib | 11.6 | .40 | .030 |
| 6 | Ponatinib | 4.2 | .41 | .517 | 6 | Ixazomib | 10.9 | .37 | .045 |
| 7 | Tepotinib | 4.1 | .41 | .518 | 7 | Trametinib | 10.4 | .73 | .019 |
| 8 | Binimetinib | 4.0 | .39 | .518 | 8 | Momelotinib | 10.3 | .37 | .034 |
| 9 | Encorafenib | 3.8 | .45 | .517 | 9 | Ruxolitinib | 9.8 | .46 | .058 |
| 10 | Pexidartinib | 3.7 | .48 | .519 | 10 | Binimetinib | 9.5 | .41 | .032 |
| *Multiple extremes total: 121 drugs (7.4%)* | | | | | *Multiple extremes total: 168 drugs (10.3%)* | | | | |

### 18 Shared Deprioritized Drugs Across Cohorts

Table 12 lists the 18 drugs deprioritized in both the cognitive and respiratory cohorts. All shared drugs showed improved S in the respiratory cohort ($\Delta$S = +0.06 to +0.16) but reduced E ($\Delta$E = $-0.38$ to $-0.44$), indicating convergent deprioritization from different failure modes.

Table 12: **Drugs deprioritized in both cohorts (n=18).** All shared drugs show improved S but reduced E in the respiratory cohort, indicating convergent deprioritization from different failure modes: S driven in the cognitive cohort, E driven in the respiratory cohort.

| | **NCT04809974** | | **NCT04880161** | | **Δ (Resp–Cog)** | |
|---|---|---|---|---|---|---|
| **Drug** | **S** | **E** | **S** | **E** | **ΔS** | **ΔE** |
| Secnidazole | 0.124 | 0.516 | 0.285 | 0.122 | +0.16 | −0.39 |
| Ethionamide | 0.196 | 0.517 | 0.306 | 0.129 | +0.11 | −0.39 |
| Tavaborole | 0.168 | 0.519 | 0.309 | 0.119 | +0.14 | −0.40 |
| Isoflurophate | 0.240 | 0.517 | 0.347 | 0.109 | +0.11 | −0.41 |
| Isradipine | 0.220 | 0.517 | 0.340 | 0.124 | +0.12 | −0.39 |
| Butoconazole | 0.169 | 0.519 | 0.303 | 0.131 | +0.13 | −0.39 |
| Lincomycin | 0.234 | 0.518 | 0.342 | 0.123 | +0.11 | −0.40 |
| Hydroxocobalamin | 0.191 | 0.520 | 0.329 | 0.116 | +0.14 | −0.40 |
| Uridine triacetate | 0.243 | 0.518 | 0.318 | 0.134 | +0.08 | −0.38 |
| Olmutinib | 0.273 | 0.518 | 0.345 | 0.116 | +0.07 | −0.40 |
| Canagliflozin | 0.196 | 0.518 | 0.330 | 0.123 | +0.13 | −0.40 |
| Ethosuximide | 0.228 | 0.518 | 0.343 | 0.115 | +0.12 | −0.40 |
| Linagliptin | 0.231 | 0.518 | 0.357 | 0.109 | +0.13 | −0.41 |
| Capmatinib | 0.312 | 0.518 | 0.380 | 0.108 | +0.07 | −0.41 |
| Phenazopyridine | 0.257 | 0.517 | 0.350 | 0.117 | +0.09 | −0.40 |
| Permethrin | 0.242 | 0.519 | 0.298 | 0.076 | +0.06 | −0.44 |
| Entrectinib | 0.324 | 0.519 | 0.403 | 0.088 | +0.08 | −0.43 |
| Copanlisib | 0.341 | 0.519 | 0.416 | 0.085 | +0.08 | −0.43 |

### 19 Enrichment Analysis — Full Results

In the cognitive cohort, the avoid list is enriched for agents with FDA black box warnings (JAK inhibitors: upadacitinib, filgotinib, tofacitinib), documented hepatotoxicity signals (secnidazole, ethionamide, fidaxomicin [12]), and oncology kinase inhibitors with multi-system toxicity profiles (cobimetinib, regorafenib, sunitinib [5, 1]). In the respiratory cohort, the avoid list is enriched for cancer agents with myelosuppressive, nephrotoxic, and neurotoxic profiles (pazopanib, azacitidine, carboplatin, paclitaxel [19, 18, 20]), as well as cholinergic agents whose AE profile may be amplified in patients with autonomic instability (donepezil, S=0.279 [14]).

Of the 18 drugs currently under active investigation in Long COVID clinical trials that appear on the SPLIT avoid lists, nine were flagged in both cohorts (acetylcysteine, acetylsalicylic acid, apixaban, baricitinib, cholecalciferol, ergocalciferol, ivermectin, ritonavir, upadacitinib), and the remainder were cohort-specific. The primary avoidance triggers, cohort assignments, study types, and supporting references for each drug are summarized in the main manuscript.

### 20 Supplementary Figures

The following figures provide extended visual support for the results reported in the main manuscript. Figure 3 shows the full S versus E scatter for all 1,639 screened drugs alongside composite score distributions with deprioritization thresholds marked, giving a complete view of how the panel is structured in each cohort. Figure 4 shows the score distributions within the deprioritized subset specifically, quantifying the failure profiles of flagged drugs beyond what the full-panel distributions reveal. Figure 5 extends the cross-cohort analysis by showing the complete ΔS versus ΔE scatter for all 18 shared deprioritized drugs and connected line plots for all 16 cross-phenotype shortlist candidates, complementing the seven-drug dumbbell plot shown in Figure 7 of the main manuscript.

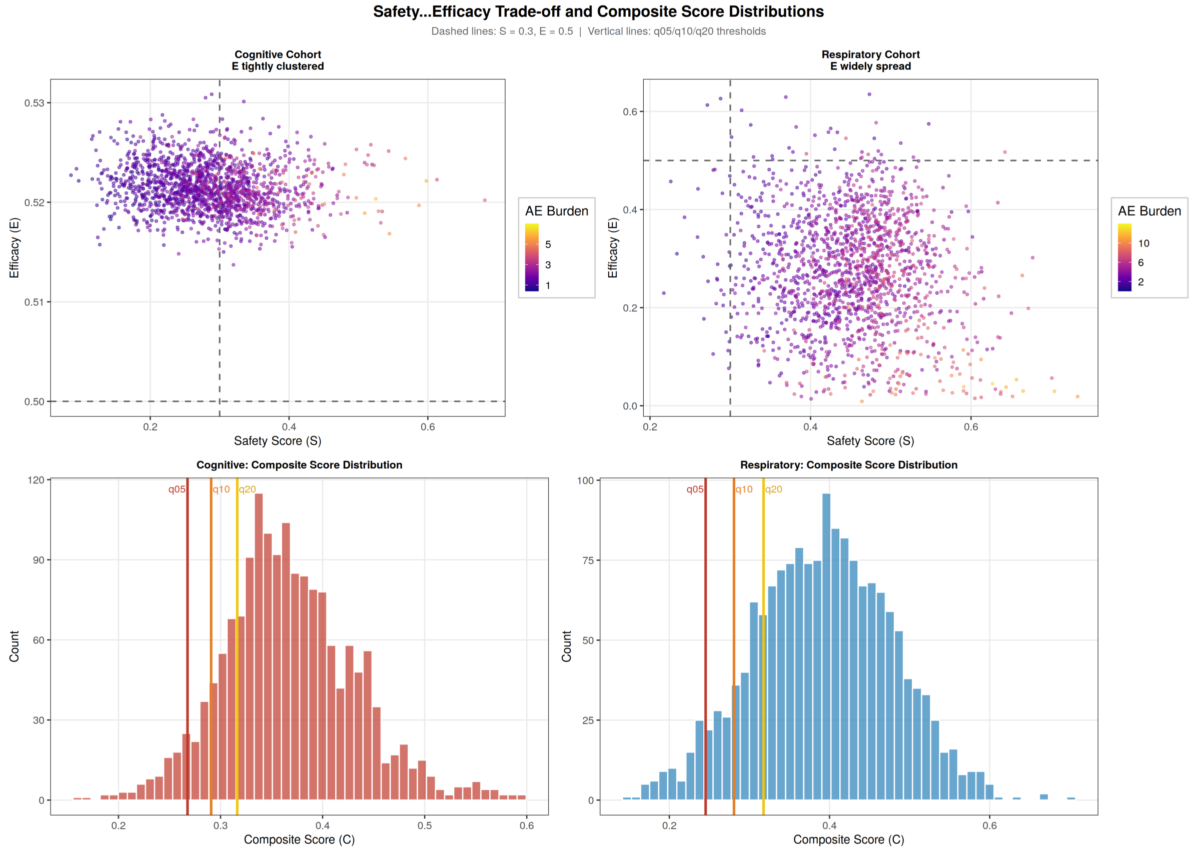

Figure 3: **Full-panel S versus E scatter and composite score distributions with deprioritization thresholds.** Upper panels: scatter plots of predicted S against predicted E for all 1,639 screened drugs in the cognitive (left) and respiratory (right) cohorts, with AE burden encoded as point color. Dashed lines indicate S=0.3 and E=0.5 reference thresholds. Lower panels: histograms of composite score $C$ for each cohort, with vertical lines marking the q05 (red), q10 (orange), and q20 (yellow) deprioritization thresholds. The cognitive cohort distribution is left-skewed with a long lower tail; the respiratory cohort is more symmetric and shifted right, reflecting higher baseline S in that context. **Abbreviations:** S, predicted safety score; E, predicted comparative efficacy score; AE, adverse event burden score; C, composite score.

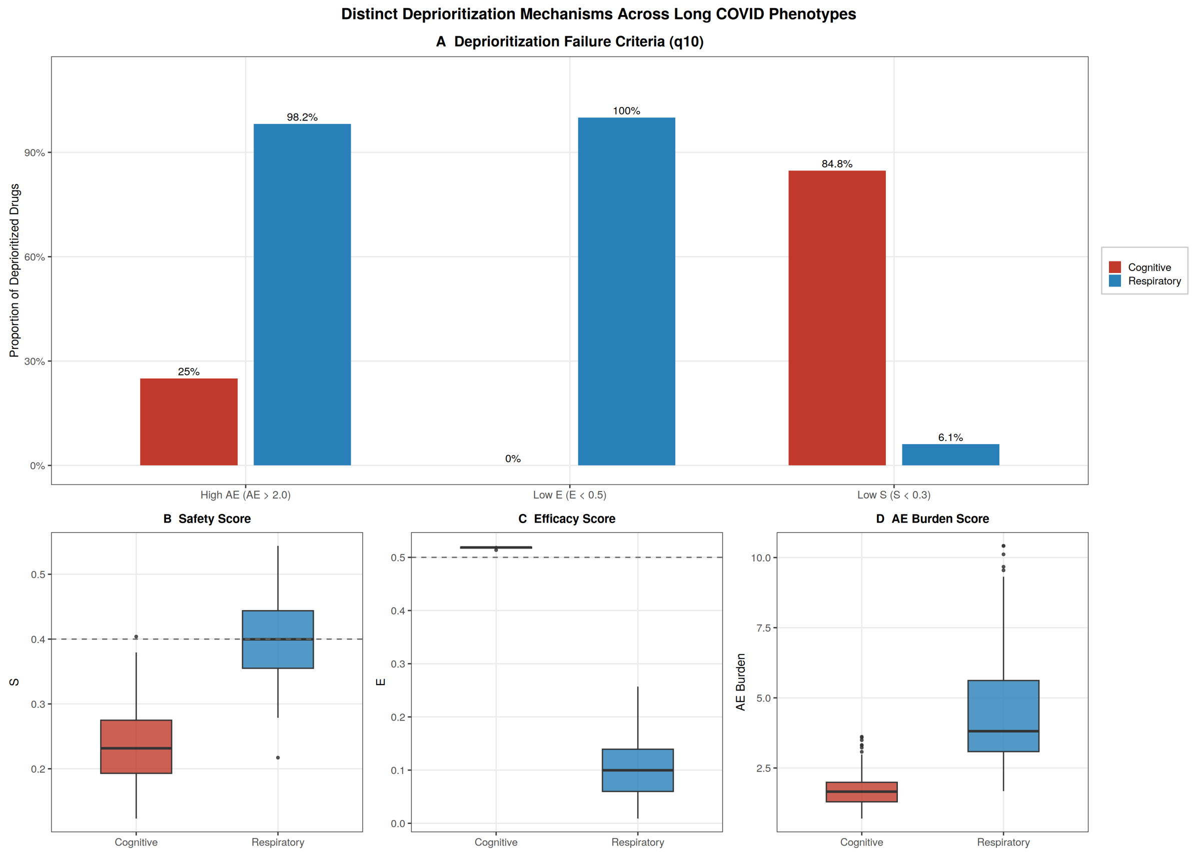

Figure 4: **Score distributions within the deprioritized subset across cohorts.** (A) Proportion of deprioritized drugs (q10) meeting each failure criterion: high AE burden (AE>2.0), low E (E<0.5), and low S (S<0.3). The cognitive cohort is dominated by low S (84.8%); the respiratory cohort by low E (100%) and high AE (98.2%). (B–D) Box plots of S, E, and AE burden scores within the deprioritized q10 subset for both cohorts. The cognitive deprioritized subset has lower S (median ≈0.23) but E compressed near placebo; the respiratory deprioritized subset has near-zero E (median ≈0.09) but higher S and substantially elevated AE burden. Dashed lines: S=0.4 (panel B) and E=0.5 (panel C). **Abbreviations:** S, predicted safety score; E, predicted comparative efficacy score; AE, adverse event burden score.

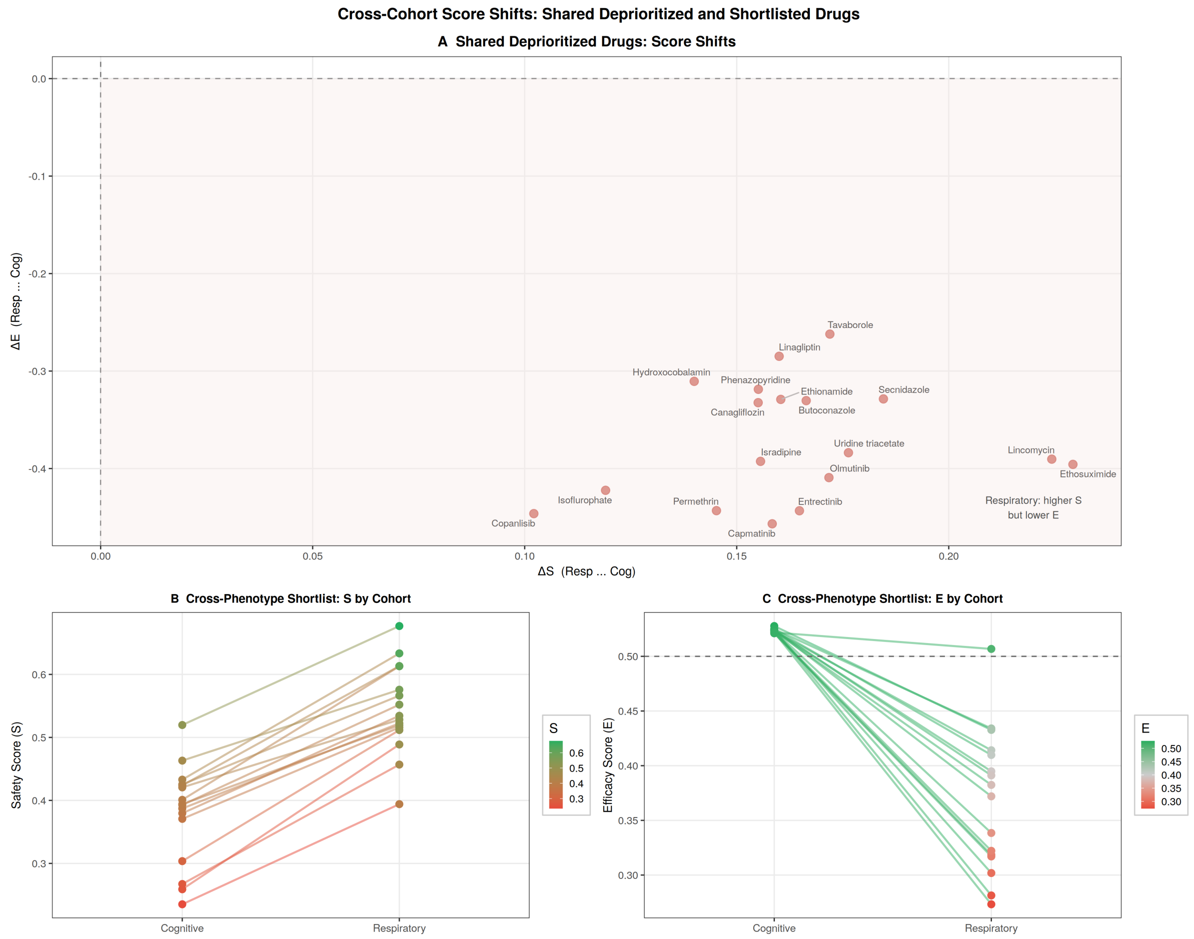

Figure 5: **Cross-cohort score shifts for shared deprioritized drugs and cross-phenotype shortlist candidates.** (A) Scatter plot of ΔS versus ΔE (respiratory minus cognitive) for the 18 drugs deprioritized in both cohorts. All drugs fall in the upper-left quadrant (ΔS>0, ΔE<0), indicating that moving to the respiratory context improves predicted safety but reduces predicted efficacy. (B) Connected line plot of S by cohort for the 16 cross-phenotype shortlist candidates. Color encodes cognitive S value; lines show the direction and magnitude of the S shift. (C) Connected line plot of E by cohort for the same candidates. Dashed line at E=0.5 marks placebo equivalence. The majority of candidates show declining E in the respiratory context, consistent with the broader efficacy compression observed in that cohort. **Abbreviations:** S, predicted safety score; E, predicted comparative efficacy score; Δ, respiratory minus cognitive difference.

### References

[1] Omar Abdel-Rahman and Mohamed Fouad. Risk of cardiovascular toxicities in patients with solid tumors treated with sunitinib, axitinib, cediranib or regorafenib: An updated systematic review and comparative meta-analysis. *Critical Reviews in Oncology/Hematology*, 92(3):194–207, 2014.

[2] Matko Brbic, Michihiro Yasunaga, Pranav Agarwal, and Jure Leskovec. Planet: Predicting population response to drugs via clinical knowledge graph. *medRxiv*, 2024.

[3] Seth Falcon and Robert Gentleman. Using GOstats to test gene lists for GO term association. *Bioinformatics*, 23(2):257–258, 2007.

[4] Anna Gaulton et al. Chembl: a large-scale bioactivity database for drug discovery. *Nucleic Acids Research*, 40(D1):D1100–D1107, 2012.

[5] Pooja Ghatalia, Youjin Je, Mehmet D. Kaymakcalan, Guru P. Sonpavde, and Toni K. Choueiri. Hepatotoxicity with vascular endothelial growth factor receptor tyrosine kinase inhibitors: A meta-analysis of randomized clinical trials. *Critical Reviews in Oncology/Hematology*, 93(3):257–276, 2015.

[6] K. J. Gleason, F. Yang, and L. S. Chen. A robust two-sample transcriptome-wide Mendelian Randomization method integrating GWAS with multi-tissue eQTL summary statistics. *Genetic Epidemiology*, 45(4):353–371, Jun 2021.

[7] K. P. Jablonski, M. Pirkl, D. Ćevid, P. Bühlmann, and N. Beerenwinkel. Identifying cancer pathway dysregulations using differential causal effects. *Bioinformatics*, 38(6):1550–1559, 2022.

[8] Sunghwan Kim, Paul A. Thiessen, Evan E. Bolton, and Stephen H. Bryant. Pubchem pug-rest: a restful web service for programmatic access to pubchem data. *Nucleic Acids Research*, 46(W1):W563–W570, 2018.

[9] Sunghwan Kim, Paul A. Thiessen, Evan E. Bolton, Jie Chen, Gang Fu, Asta Gindulyte, Lianyi Han, Jane He, Siqian He, Benjamin A. Shoemaker, Jie Wang, Bo Yu, Jian Zhang, and Stephen H. Bryant. Pubchem: a public information system for analyzing bioactivities of small molecules. *Nucleic Acids Research*, 44(D1):D1202–D1213, 2016.

[10] Christopher A. Lipinski, Franco Lombardo, Beryl W. Dominy, and Paul J. Feeney. Experimental and computational approaches to estimate solubility and permeability in drug discovery and development settings. *Advanced Drug Delivery Reviews*, 23(1–3):3–25, 1997.

[11] National Cancer Institute. Common terminology criteria for adverse events (ctcae) v6.0 (meddra 28.0).

[12] National Institute of Diabetes and Digestive and Kidney Diseases. Ethionamide, 2020. LiverTox: Clinical and Research Information on Drug-Induced Liver Injury; Accessed 27 Jan 2026.

[13] David Ochoa et al. Open targets platform: supporting systematic drug–target identification and prioritisation. *Nucleic Acids Research*, 49(D1):D1302–D1310, 2021.

[14] Laura Y. Park-Wyllie, Muhammad M. Mamdani, Philip Li, Sudeep S. Gill, Andreas Laupacis, and David N. Juurlink. Cholinesterase inhibitors and hospitalization for bradycardia: A population-based study. *PLoS Medicine*, 6(9):e1000157, 2009.

[15] Tuan Minh Pham, Sergei Rybakov, Hiroki Suzuki, Ka-Chun Yeung, Seungjin Hong, and Jie Sun. Optimal control nodes in disease-perturbed networks as targets for combination therapy. *Nature Communications*, 10:401, 2019.

[16] Sindy Pinero, Xiaomei Li, Lin Liu, Jiuyong Li, Sang Hong Lee, Marnie Winter, Thin Nguyen, Junpeng Zhang, and Thuc Duy Le. Integrative multi-omics framework for causal gene discovery in long covid. *medRxiv*, page 2025.02.09.25321751, 2025.

[17] Sindy Licette Pinero, Xiaomei Li, Lin Liu, Jiuyong Li, Sang Hong Lee, Marnie Winter, Thin Nguyen, Junpeng Zhang, and Thuc Duy Le. Taco: Tabpfn augmented causal outcomes for early detection of long covid. *medRxiv*, page 2025.10.02.25337138, 2025.

[18] U.S. Food and Drug Administration. Paraplatin (carboplatin) prescribing information, 2010. Warnings include severe myelosuppression; nephrotoxicity risk with concomitant nephrotoxins; Accessed 27 Jan 2026.

[19] U.S. Food and Drug Administration. Votrient (pazopanib) prescribing information, 2012. Boxed warning: hepatotoxicity; Accessed 27 Jan 2026.

[20] U.S. Food and Drug Administration. Paclitaxel protein-bound particles for injectable suspension (albumin-bound) prescribing information, 2023. Warnings include severe myelosuppression; peripheral neuropathy/neurotoxicity discussed in labeling; Accessed 27 Jan 2026.

[21] U.S. National Library of Medicine. Dailymed: Drug label information. National Library of Medicine (NLM), n.d. Accessed programmatically via the DailyMed services API.

[22] A. Vinayagam, T. E. Gibson, H. J. Lee, B. Yilmazel, C. Roesel, Y. Hu, Y. Kwon, A. Sharma, Y. Y. Liu, N. Perrimon, and A. L. Barabási. Controllability analysis of the directed human protein interaction network identifies disease genes and drug targets. *Proceedings of the National Academy of Sciences of the United States of America*, 113(18):4976–4981, May 2016.

[23] Hua Xu, Shane P Stenner, Son Doan, Kevin B Johnson, Lemuel R Waitman, and Joshua C Denny. MedEx: a medication information extraction system for clinical narratives. *Journal of the American Medical Informatics Association*, 17(1):19–24, 2010.

[24] Chi Yuan, Patrick B Ryan, Casey Ta, Yixuan Guo, Ziran Li, Jill Hardin, Rupa Makadia, Peng Jin, Ning Shang, Tian Kang, and Chunhua Weng. Criteria2Query: a natural language interface to clinical databases for cohort definition. *Journal of the American Medical Informatics Association*, 26(4):294–305, 2019.
